# Testing Black-White disparities in biological aging in older adults in the United States: analysis of DNA-methylation and blood-chemistry methods

**DOI:** 10.1101/2021.03.02.21252685

**Authors:** GH Graf, CL Crowe, M Kothari, D Kwon, JJ Manly, IC Turney, L Valeri, DW Belsky

**Affiliations:** Department of Epidemiology, Columbia University Mailman School of Public Health, New York, NY 10032, USA; Robert N Butler Columbia Aging Center, Columbia University Mailman School of Public Health, New York, NY 10032, USA; Psychiatric Epidemiology Training Program, Columbia University Mailman School of Public Health, New York, NY 10032, USA; Taub Institute for Research on Alzheimer’s Disease and the Aging Brain, College of Physicians and Surgeons, Columbia University, New York, NY 10032, USA; Gertrude H. Sergievsky Center, College of Physicians and Surgeons, Columbia University, New York, NY 10032, USA; Department of Neurology, College of Physicians and Surgeons, Columbia University, New York, NY 10032, USA; Department of Biostatistics, Columbia University Mailman School of Public Health, New York, NY 10032, USA

**Author notes:** **Correspondence to:** Daniel W. Belsky.

## Abstract

Biological aging is a proposed mechanism through which social determinants drive health disparities. We conducted proof-of-concept testing of eight DNA-methylation and blood-chemistry quantifications of biological aging as mediators of disparities in healthspan between Black and White participants in the United States Health and Retirement Study (HRS; n=9005). We quantified biological aging from four DNA-methylation “clocks” (Horvath, Hannum, PhenoAge, and GrimAge), a DNA-methylation Pace of Aging (DunedinPoAm), and three blood-chemistry measures (PhenoAge, Klemera-Doubal method Biological Age, and homeostatic dysregulation). We quantified Black-White disparities in healthspan from cross-sectional and longitudinal data on physical-performance tests, self-reported activities of daily living (ADL) limitations and physician-diagnosed chronic diseases, self-rated health, and survival. DNA-methylation and blood-chemistry quantifications of biological aging were moderately correlated (Pearson-r range 0.1-0.4). GrimAge, DunedinPoAm and all three blood-chemistry measures were associated with healthspan characteristics (e.g. mortality effect-size range 1.71-2.32) and showed evidence of more advanced/faster biological aging in Black compared with White participants (Cohen’s d=.4-.5). These measures accounted for 13-95% of Black-White differences in healthspan-related characteristics. Findings that Black Americans are biologically older and aging more rapidly than White Americans of the same chronological age suggest that eliminating disparities in the pace of aging can contribute building to aging health equity.

## INTRODUCTION

Black Americans experience excess morbidity and premature mortality relative to White Americans (1). This health disparity is mediated by multiple chronic diseases affecting different organ systems throughout the body and reflects an etiology extending from the earliest stages of life across adulthood, encompassing social, economic, and environmental factors (2-4). Differences in health between Black and White Americans vary between geographic locations and have changed over time, indicating that these disparities are socially determined and that they are modifiable (5-7). A range of policies and programs are proposed to mitigate health disparities (8-11). However, rigorous evaluation of impact is challenging (12). Interventions to address health disparities delivered to older adults may come too late to prevent chronic disease (4, 13), while interventions delivered to younger people require long follow-up intervals to establish impact (14). Methods are needed to monitor effectiveness of interventions over timescales of years rather than decades.

Measurements that quantify processes of biological aging may provide near-term measurements of long-term impacts. Biological aging is the gradual and progressive decline in system integrity with advancing chronological age (15). This process is now being studied as a modifiable root cause of many different chronic diseases (16-18). One hypothesis advanced to explain Black-White health disparities across a range of diseases is that social and material stresses experienced by Black Americans act to accelerate biological aging, referred to as “weathering” (19, 20). In epidemiologic studies, Black Americans show more advanced biological aging as compared to White Americans of the same chronological age (21-23). If advanced biological aging is a mediator of health disparities, then quantifications of biological aging could be used to monitor intervention impacts.

Many methods are proposed to quantify biological aging from several biological levels of analysis (24, 25). Agreement between measures is often poor; there is no gold standard (26-28). Measures based on analysis of blood-chemistry and DNA-methylation data have received the most attention to date. We conducted proof-of-concept testing of eight blood-chemistry and DNA-methylation methods to quantify biological aging as mediators of Black-White disparities in healthy aging. We analyzed deficits in physical functioning, limitations to activities of daily living, chronic disease morbidity, and mortality in a national sample of older adults in the US Health and Retirement Study (HRS). Previous studies have documented Black-White differences in several of the measures of aging we analyzed (21, 22, 29-31). However, few studies have tested if these differences could account for Black-White health disparities (21, 23) and none have specifically considered disparities in healthspan characteristics. Our analysis builds on an initial report of differences in DNA-methylation measures of biological aging between Black and White adults in the HRS (30) in three ways. First, we analyze measures of biological aging derived from DNA-methylation data together with measures derived from blood-chemistry data. Second, we compare the different measures of biological aging by testing associations with healthspan-related characteristics and mortality. Third, we quantify the fraction of Black-White differences in healthspan-related characteristics and mortality that are accounted for by biological aging measures.

## METHODS

### Sample

The Health and Retirement Study (HRS) is a nationally representative longitudinal survey of US residents ≥50 years of age and their spouses fielded every two years since 1992. A new cohort of 51-56 year-olds and their spouses is enrolled every six years to maintain representativeness of the U.S. population over 50 years of age. Response rates over all waves of the HRS range from 81-91%. As of the most recent data release, HRS included data collected from 42,515 individuals in 26,600 households. We linked HRS data curated by RAND Corporation with new data collected as part of HRS’s 2016 Venous Blood Study. Analysis included participants aged 50-90 years at the time of the blood draw for whom data were also collected on prevalent chronic disease, limitations to activities of daily living, and/or testing to assess physical functioning (n=9005). Comparison of our analysis sample to the full HRS is reported in Supplemental Table S1.

### Measures

#### Biological Aging

There is no gold standard measure of biological aging (24). Many methods have been proposed based on different biological levels of analysis. Current state-of-the-art methods use machine learning to sift large numbers of candidate markers and parameterize algorithms that predict aging-related parameters, including chronological age, mortality risk, and rate of decline in system integrity. Algorithms are developed in reference datasets and applied in new datasets to test hypotheses. We analyzed several different methods because each method uses different assumptions to develop a measure of the latent construct of biological aging. As we and others have shown, the different methods do not all measure the same aspects of the aging process (26, 28). Comparative analysis is therefore essential to interpretation. We focused on algorithms developed for blood-chemistry analytes routinely measured in clinical settings and DNA-methylation marks included on commercial arrays, and which have received substantial attention in the research literature. Measures are summarized and their source publications are cited in **Table 1**. Detailed descriptions of the measures are reported in the **Supplemental Methods Section 1**.

**Table 1.** Measures of Biological Aging Included in Analysis. The table reports the eight measures of biological aging included in analysis. For each measure, the table reports the criterion used to develop the measure and the interpretation of the measure’s values.

|  | Criterion | Interpretation |
| --- | --- | --- |
| **Blood Chemistry Measures.** All algorithms were parameterized using data from NHANES III and included the following blood chemistries: albumin, alkaline phosphatase, creatinine, C-reactive protein (log), glucose, white blood cell count, lymphocyte %, mean cell volume, and red cell distribution width. PhenoAge and KDM Biological Age algorithms additionally included chronological age. The NHANES III training sample is majority White, but includes an oversample of Black Americans. Blood chemistry measures were calculated using code available on GitHub (https://rdrr.io/github/dayoonkwon/BioAge/) according to published methods. For analysis, PhenoAge and KDM Biological Age were differenced from chronological age to calculate biological-age advancement values. |  |  |
| PhenoAge (50) | Mortality | Age at which average mortality risk in NHANES III matches the mortality risk predicted by the blood-chemistry + chronological age algorithm. |
| KDM Biological Age (51) | Chronological Age | Age at which average physiology in NHANES III matches the physiology of the participant. |
| Homeostatic Dysregulation (52) | Deviation from healthy youth | Log biomarker-Mahalanobis distance of participant from young, healthy NHANES III participants |
| **DNA-Methylation Measures.** DNA-methylation measures were developed from analysis of genome-wide DNA-methylation measured on Illumina 27k and 450k arrays in a range of different datasets. The Horvath Clock was developed from analysis of 82 different datasets. The Hannum Clock was developed from analysis of research volunteers at UC San Diego, University of Southern California, and West China Hospital. The PhenoAge Clock was developed from analysis of NHANES III and the InCHIANTI Study. The GrimAge clock was developed from analysis of the Framingham Heart Study Offspring Cohort. The DunedinPoAm Pace of Aging was developed from analysis of the Dunedin Study. The InCHIANTI, Framingham, and Dunedin cohorts are mostly or entirely White-European. DNA-methylation measures were calculated by the HRS investigators. For analysis, DNA-methylation clocks were residualized on chronological age to calculate biological-age advancement values. |  |  |
| **First Generation DNA-methylation Clocks** |  |  |
| Horvath Clock (53) | Chronological Age | Age predicted by DNA methylation. |
| Hannum Clock (54) | Chronological Age | Age predicted by DNA methylation. |
| **Second Generation DNA-Methylation Clocks** |  |  |
| PhenoAge Clock (50) | Blood-chemistry PhenoAge | Age at which average mortality risk in NHANES III matches the mortality risk predicted by the PhenoAge algorithm. |
| GrimAge Clock (41) | Mortality | Age at which average mortality risk in the Framingham Heart Study Offspring cohort matches predicted mortality risk. |
| **Pace of Aging** |  |  |
| DunedinPoAm Pace of Aging (43) | Change over 12-years of follow-up in 18 system-integrity biomarkers | Years of physiological decline experienced per 1 year of calendar time over the recent past. The expected value of DunedinPoAm in midlife adults is 1. Values>1 indicate accelerated aging. Values<1 indicate slowed aging. |

We computed blood-chemistry measures using the R package *BioAge* (https://github.com/dayoonkwon/BioAge). We obtained data on DNA-methylation measures of biological age from the HRS (30). Clock measures can also be computed using the software hosted by the Horvath Lab (http://dnamage.genetics.ucla.edu/). DunedinPoAm pace of aging can be computed using the GitHub code https://github.com/danbelsky/DunedinPoAm38.

For analysis, we converted measures of biological age (blood-chemistry PhenoAge, Klemera-Doubal Biological Age, Homeostatic Dysregulation, PhenoAge Clock, Horvath Clock, Hannum Clock, GrimAge, DunedinPoAm Pace of Aging) to measures of biological-age advancement by fitting regressions of biological age measures on chronological age and computing residual values. The literature commonly refers to these residuals as ‘age acceleration’. We instead use ‘age advancement’ to distinguish measurements like the clocks-which compute a difference between biological age and chronological age at a single point in time-from pace of aging measurements that quantify how fast a person is aging. No residualization was applied to homeostatic dysregulation and DunedinPoAm, as these measures already quantify deviation from the expected sample norm.

#### Healthspan-related characteristics

Healthspan is the portion of life lived free of disease and disability. We measured healthspan-related characteristics from performance-test measurement of functional impairment administered by trained interviewers, participant reports about activities of daily living (ADL) limitations, self-rated health, and physician-diagnosed chronic conditions, and HRS follow-up for mortality status through 2019. Measures are described in detail in **Table 2** and **Supplemental Methods Table 1**.

**Table 2.**
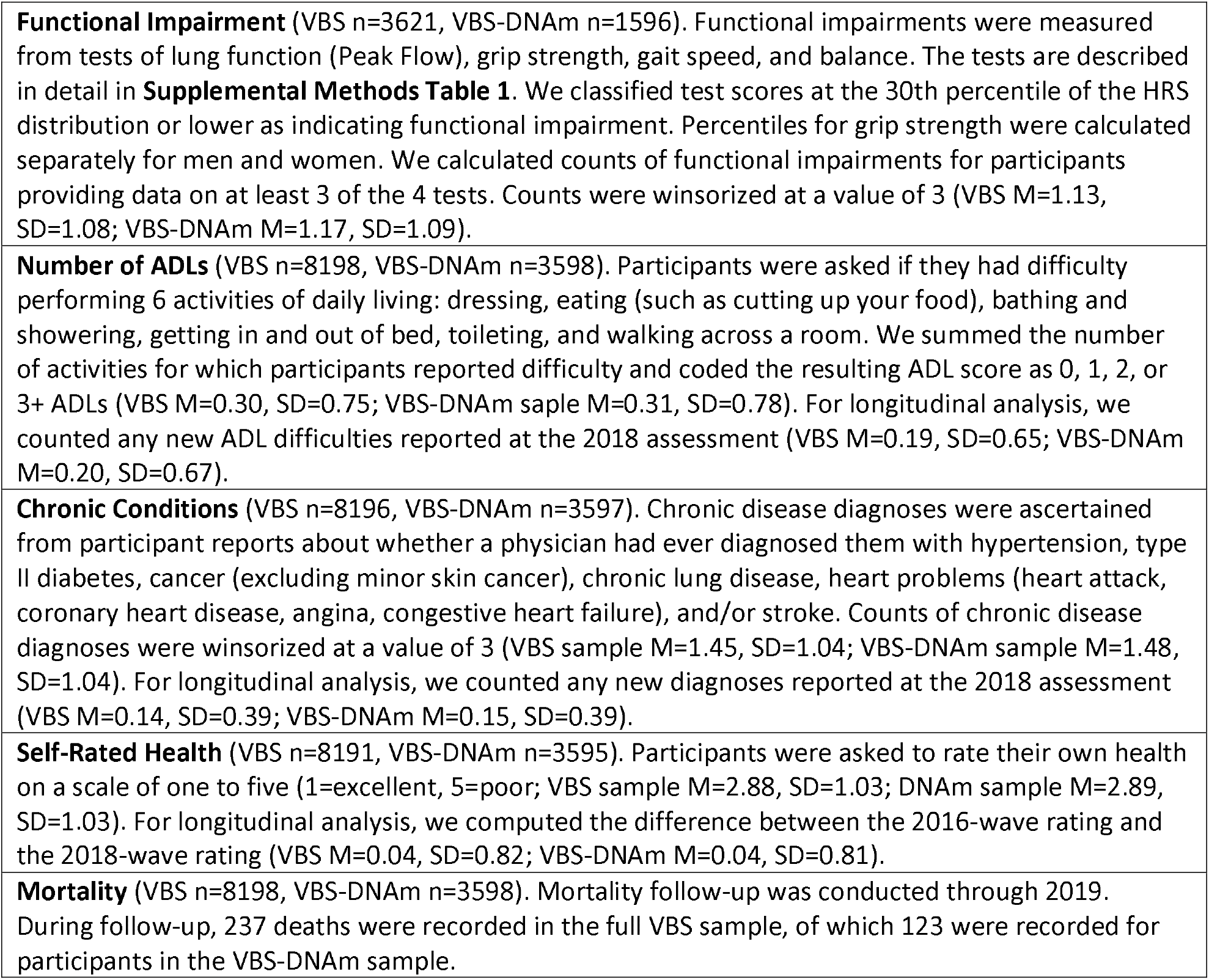
Measurement of healthspan-related characteristics and mortality.

### Analysis

Our primary analysis tested associations of biological aging measures with healthspan-related characteristics. We conducted analysis of prevalent functional impairments, ADLs, chronic conditions, and current self-rated health based on data collected in HRS’s 2016 wave. We conducted longitudinal analysis of incident ADLs, incident chronic conditions, and changes in self-rated health using data from the 2016 and 2018 waves. (We did not conduct longitudinal analysis of performance-test measures because these are collected from participants at every-other wave, so no follow-up was available). Analysis of mortality was based on the most-recent mortality status ascertainment by HRS.

We used Poisson regression to estimate Incident Rate Ratios (IRRs) for associations of biological aging with counts of functional impairments, ADLs, and chronic conditions. We used linear regression to estimate standardized effect-sizes (Pearson’s r) for continuous measures of self-rated health. We used Cox proportional hazards regression to estimate mortality hazard ratios (HR). Effect-sizes for biological aging measures were denominated in standard-deviation units (SDs).

For analysis of the full HRS VBS and VBS-DNAm samples, which were designed to represent the US population aged 50 and older, we applied probability sampling weights to generate estimates for this population. For analyses of subsamples of Black and White older adults, we included covariate adjustment for chronological age, sex, and region of residence.

We tested mediation of Black-White disparities in biological aging using a regression-based approach as described by Valeri and VanderWeele (32). We used the R package CMAverse (33) to estimate direct and indirect effects and proportions mediated. We calculated confidence intervals using bootstrapping to obtain standard error estimates. We tested robustness of mediation results to potential exposure-mediator interactions following the approaches outlined by Valeri and VanderWeele (32, 34). Details of this analysis are in **Supplemental Methods Sections 2 and 3**.

## RESULTS

We conducted two sets of analyses. First, we analyzed blood-chemistry measures of biological aging. Next, we analyzed DNA-methylation measures of biological aging, including comparison to blood-chemistry measures. Within each analysis set, we first tested associations of biological-aging measures with healthspan-related characteristics. Next, we tested Black-White disparities in biological aging. Finally, we conducted mediation analysis for those measures demonstrating associations of more-advanced/faster biological aging with healthspan-related characteristics and more-advanced/faster biological aging in Black as compared to White participants (35).

### I. Blood Chemistry Analysis

We analyzed DNA-methylation measures of aging in n=9005 HRS participants included in the Venous Blood Study (VBS; 41% male, 74% White, 18% Black, aged 50-90 years, mean age=69, SD=9). Participants’ Phenotypic Ages were highly correlated with their chronological ages (r=0.76). Results were similar for the Klemera-Doubal method (KDM) Biological Age and homeostatic dysregulation measures **(Supplemental Table S1)**.

#### Older adults with more advanced biological aging in blood-chemistry analysis showed deficits in healthspan-related characteristics and were at increased risk of mortality

Participants with more advanced biological aging measured from blood chemistries had poorer outcomes for all healthspan-related characteristics and increased risk for mortality. For Phenotypic Age advancement, increased risk for prevalent functional impairments, prevalent and incident ADLs, and prevalent and incident chronic conditions ranged from 19-54% per SD; effect-sizes were r=0.34 for current self-rated health and 0.09 for change in self-rated health; the increase in the hazard of mortality was 85% per SD. Results were similar in analysis of KDM Biological Age and homeostatic dysregulation measures (**Figure 1, Supplemental Table S2**).

**Figure 1.**
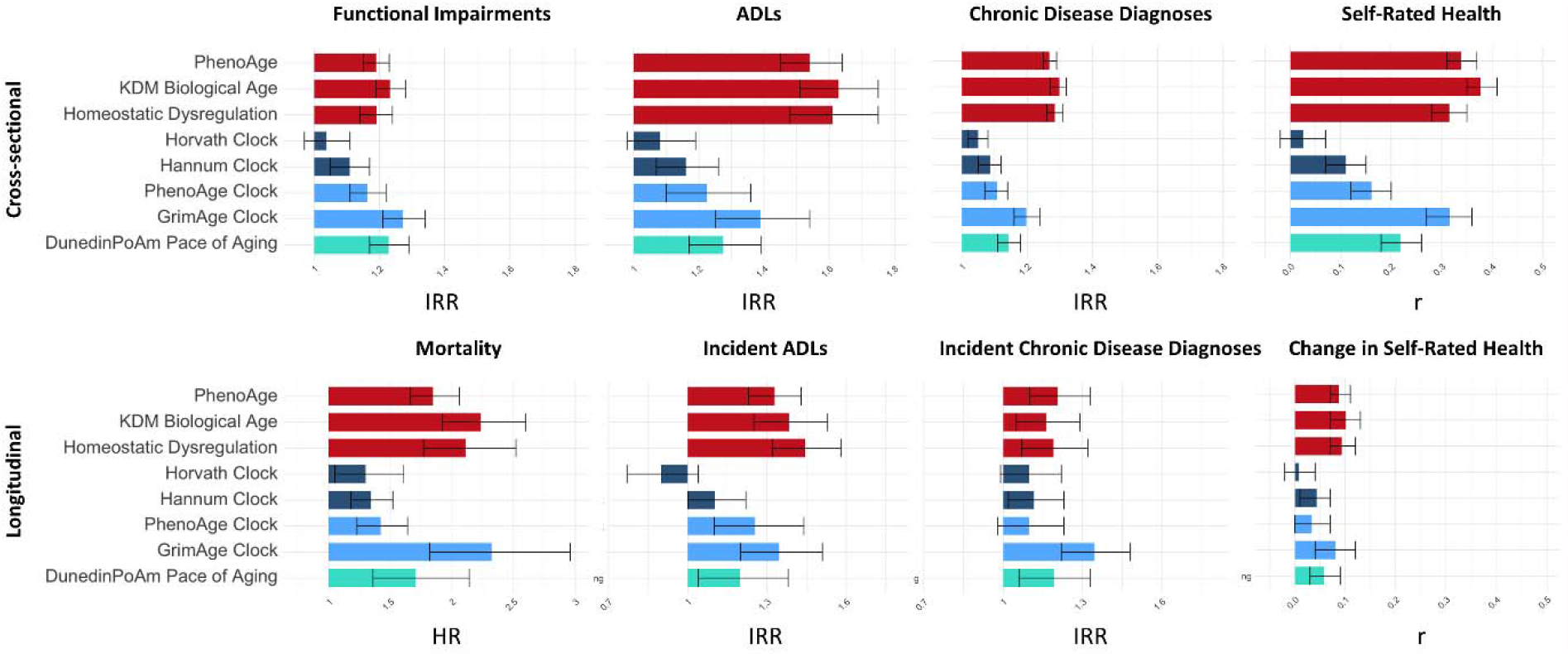
Effect-sizes for associations of biological aging measures with healthspan-related characteristics. The figure graphs effect-sizes for associations of biological aging measures with healthspan characteristics measured cross-sectionally at the 2016 wave (Functional Impairments, Limitations to Activities of Daily Living (ADLs), Chronic Disease Diagnoses, and Self-rated Health) and with healthspan-related characteristics and mortality measured longitudinally through the end of the 2018 wave (mortality, incident ADLs, incident chronic disease diagnoses, change in self-rated health). For functional impairment, prevalent and incident chronic conditions, and prevalent and incident ADLs, effect-sizes are incidence rate ratios (IRRs) for 1-SD increases in biological aging measures estimated from Poisson regression. For cross-sectional self-rated health and change in self-rated health, effect-sizes are Pearson r’s for 1-SD increases in biological aging measures estimated from linear regression. For mortality, effect-sizes are hazard ratios (HRs) for 1-SD increases in biological aging measures estimated from Poisson regression. Blood-chemistry measures are shown in red; 1st generation DNA-methylation clocks are shown in dark blue; 2nd generation DNA-methylation clocks are shown in light blue; DunedinPoAm pace of aging is shown in turquoise. Error bars show 95% confidence intervals. For cross-sectional measures, sample sizes for VBS/VBS-DNAm samples are n=3721/1676 for functional impairment; n=8484/3785 for ADLs; n=8482/3784 for chronic disease diagnoses; n=8476/3782 for self-rated health. For longitudinal measures, sample sizes for VBS/VBS-DNAm samples are n=8484/3785 for mortality; n=7491/3327 for ADLs; n=7497/3329 for chronic conditions; n=7488/3326 for self-rated health.

#### Black HRS VBS participants showed deficits in healthspan-related characteristics and increased risk for mortality as compared with White participants

Black participants more often demonstrated functional impairments (IRR=1.25 [1.15-1.35]), and reported more ADLs (IRR=1.91 [1.75-2.10]) and diagnosed chronic conditions (IRR=1.26 [1.20-1.32]) and poorer self-rated health (d=0.33 [0.28-0.39]) as compared with White participants. Over follow-up, Black participants were more likely to report incident ADLs (IRR=1.49 [1.31-1.69]) and declines in self-rated health (d=0.11 [0.06-0.15]). Black-White differences in mortality risk were in the expected direction but not statistically different from zero at the alpha=0.05 threshold (HR=1.36 [0.97-1.91]). Black-White differences were in the opposite direction for incident chronic conditions (IRR=0.84 [0.64-1.10]), possibly reflecting differences in access to care resulting in under-diagnosis among Black participants and/or the overall higher burden of chronic disease prevalent at baseline in the Black as compared to White participants. Effect-sizes are reported in **Supplemental Table S3**.

#### Black HRS VBS participants showed more advanced biological aging in blood-chemistry analysis as compared with White participants

Black participants experienced an additional 2.93 years of biological aging (95% CI [2.47-3.40]) as compared to White participants of the same chronological age, according to the blood-chemistry PhenoAge measure. Results were similar for analysis of KDM Biological Age and homeostatic dysregulation (**Figure 2, Supplemental Table S1**).

**Figure 2.**
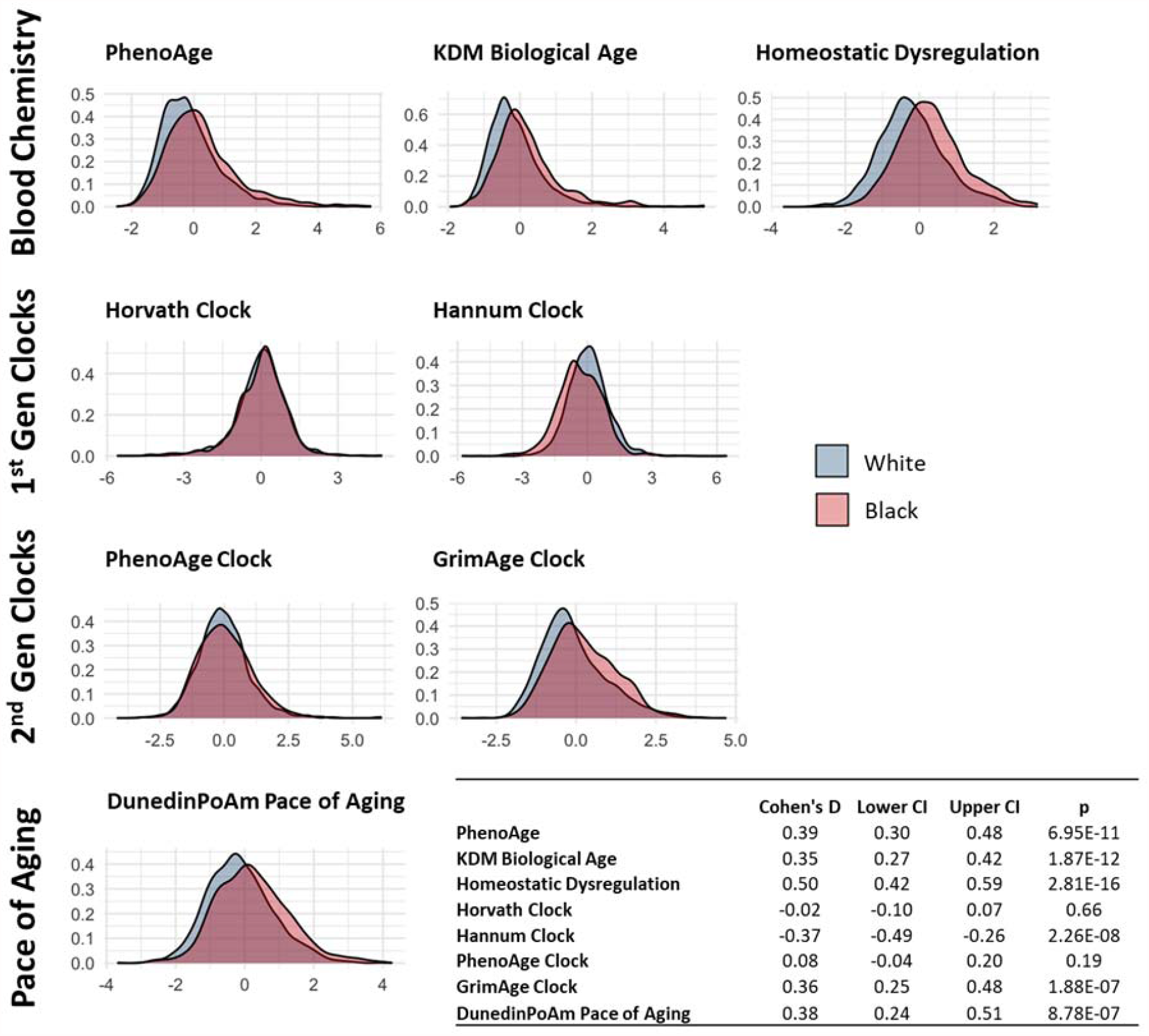
Distribution of biological aging measures in Black and White US Health and Retirement Study participants. The figure graphs densities of biological aging measures. For blood-chemistry PhenoAge, KDM Biological Age, and the DNA-methylation clocks, values are biological-age advancements (i.e. the difference between measured biological age and chronological age). For homeostatic dysregulation, values capture blood-chemistry deviation from the norm in a healthy sample. For DunedinPoAm, values are pace of aging (i.e. years of physiological decline experienced per 1 year of calendar time over the recent past). To allow comparison across measures, biological aging values are standardized to mean=0, SD=1 in the full VBS sample. Densities and Cohen’s D estimates are adjusted for sampling weights.

#### Blood-chemistry quantifications of biological aging accounted for significant proportions of Black-White differences in healthspan-related characteristics and mortality risk

More advanced blood-chemistry PhenoAge accounted for 20-30% of Black-White differences in healthspan-related characteristics and more than half of the Black-White difference in mortality risk (proportion mediated=0.19 for functional impairments, 0.18 for prevalent ADLs, 0.17 for incident ADLs, 0.29 for chronic conditions, 0.29 for self-rated health; 0.18 for changes in self-rated health, and 0.60 for mortality; we did not compute a mediation proportion for incident chronic conditions because incidence was higher in White as compared with Black participants). Results were similar for analysis of KDM Biological Age and homeostatic dysregulation, although the proportions mediated were somewhat larger for the homeostatic dysregulation measure (range=0.27-0.93). Effect-size estimates for Black-White differences in healthspan-related characteristics before and after adjustment for biological aging measures are reported in **Supplemental Table S3**. Mediation analysis results are reported in **Table 3** and **Supplemental Table 4**.

**Table 3.** Mediation analysis of Black-White disparities in healthspan-related characteristics. The table shows parameter estimates and proportion-mediation calculations from mediation analysis. Results are presented for analysis of different healthspan-related characteristics and mortality in the table rows. For each outcome, there are three columns of results corresponding to the PhenoAge blood chemistry measure (left), the GrimAge DNA methylation clock (center), and the DunedinPoAm pace of aging (right). For each outcome-aging measure pair, there are two mediation analyses. The first analysis (“Without E-M” interaction”) estimates the mediation model under the assumption that associations between biological aging measures and healthspan-related characteristics/mortality are the same in the Black and White subsamples (i.e. no exposure-mediator interaction). The second analysis (“With E-M Interaction” estimates the mediation model allowing for different associations between biological aging measures and healthspan-related characteristics in the Black and White subsamples (i.e. with explicit modeling of exposure-mediator interaction). Indirect effects estimates represent the portion of the Black-White disparity mediated through the biological aging measure. Total effect estimates represent the Black-White healthspan disparity. The proportion mediated is computed as a ratio of the indirect to the total effect. **Panel A** shows results for mediation analysis of cross-sectional data collected in the 2016 wave of the HRS. Samples sizes for VBS/VBS-DNAm samples are n=3621/1596 for functional impairment; n=8197/3598 for ADLs; n=8195/3597 for chronic conditions; n=8190/3595 for self-rated health. **Panel B** shows results for mediation analysis of longitudinal data in which measures of biological aging were collected in the 2016 wave and mortality, incident ADLs and chronic conditions, and changes in self-rated health were measured from 2016 baseline through follow-up in the 2018 data collection wave. Sample sizes for VBS/VBS-DNAm samples are n=8197/3598 for mortality; n=8197/3598 for ADLs; n=7229/3164 for chronic conditions; n=7228/3163 for self-rated health.

| Panel A. Cross-sectional Measures |  |  |  |  |  |  |  |  |  |  |  |  |
| --- | --- | --- | --- | --- | --- | --- | --- | --- | --- | --- | --- | --- |
|  | PhenoAge |  |  |  | GrimAge |  |  |  | DunedinPoAm |  |  |  |
|  | Without E-M interaction |  | With E-M interaction |  | Without E-M interaction |  | With E-M interaction |  | Without E-M interaction |  | With E-M interaction |  |
|  | Est | 95%CI | Est | 95%CI | Est | 95%CI | Est | 95%CI | Est | 95%CI | Est | 95%CI |
| **Functional Impairment** |  |  |  |  |  |  |  |  |  |  |  |  |
| Indirect effect | 0.04 | ( 0.03, 0.05) | 0.04 | ( 0.02, 0.06) | 0.08 | ( 0.06, 0.10) | 0.08 | ( 0.04, 0.12) | 0.06 | ( 0.04, 0.08) | 0.04 | ( 0.00, 0.07) |
| Total effect | 0.22 | ( 0.15, 0.29) | 0.22 | ( 0.15, 0.28) | 0.18 | ( 0.09, 0.31) | 0.18 | ( 0.08, 0.31) | 0.17 | ( 0.06, 0.29) | 0.17 | ( 0.08, 0.28) |
| Prop. Mediated | 0.19 |  | 0.17 |  | 0.43 |  | 0.42 |  | 0.34 |  | 0.23 |  |
| **ADLs** |  |  |  |  |  |  |  |  |  |  |  |  |
| Indirect effect | 0.12 | ( 0.09, 0.14) | 0.07 | ( 0.05, 0.10) | 0.10 | ( 0.07, 0.14) | 0.05 | ( 0.01, 0.11) | 0.08 | ( 0.05, 0.11) | 0.04 | ( -0.01, 0.08) |
| Total effect | 0.63 | ( 0.52, 0.74) | 0.66 | ( 0.55, 0.79) | 0.59 | ( 0.37, 0.80) | 0.58 | ( 0.40, 0.75) | 0.59 | ( 0.41, 0.75) | 0.59 | ( 0.42, 0.77) |
| Prop. Mediated | 0.18 |  | 0.11 |  | 0.16 |  | 0.09 |  | 0.13 |  | 0.06 |  |
| **Chronic Conditions** |  |  |  |  |  |  |  |  |  |  |  |  |
| Indirect effect | 0.07 | ( 0.05, 0.08) | 0.05 | ( 0.04, 0.07) | 0.06 | ( 0.04, 0.07) | 0.03 | ( 0.02, 0.05) | 0.04 | ( 0.03, 0.06) | 0.02 | ( 0.01, 0.04) |
| Total effect | 0.23 | ( 0.19, 0.25) | 0.23 | ( 0.20, 0.27) | 0.22 | ( 0.16, 0.27) | 0.22 | ( 0.16, 0.28) | 0.22 | ( 0.16, 0.28) | 0.22 | ( 0.16, 0.28) |
| Prop. Mediated | 0.29 |  | 0.23 |  | 0.26 |  | 0.14 |  | 0.19 |  | 0.09 |  |
| **Self-Rated Health** |  |  |  |  |  |  |  |  |  |  |  |  |
| Indirect effect | 0.10 | ( 0.08, 0.12) | 0.07 | ( 0.06, 0.10) | 0.10 | ( 0.08, 0.13) | 0.10 | ( 0.07, 0.13) | 0.07 | ( 0.05, 0.09) | 0.08 | ( 0.06, 0.12) |
| Total effect | 0.33 | ( 0.27, 0.40) | 0.34 | ( 0.28, 0.39) | 0.28 | ( 0.20, 0.37) | 0.28 | ( 0.20, 0.35) | 0.28 | ( 0.20, 0.37) | 0.28 | ( 0.20, 0.36) |
| Prop. Mediated | 0.29 |  | 0.22 |  | 0.37 |  | 0.35 |  | 0.24 |  | 0.29 |  |
| Panel B. Longitudinal Measures |  |  |  |  |  |  |  |  |  |  |  |  |
|  | PhenoAge |  |  |  | GrimAge |  |  |  | DunedinPoAm |  |  |  |
|  | Without E-M interaction |  | With E-M interaction |  | Without E-M interaction |  | With E-M interaction |  | Without E-M interaction |  | With E-M interaction |  |
|  | Est | 95%CI | Est | 95%CI | Est | 95%CI | Est | 95%CI | Est | 95%CI | Est | 95%CI |
| **Mortality** |  |  |  |  |  |  |  |  |  |  |  |  |
| Indirect effect | 0.16 | ( 0.12, 0.21) | 0.16 | ( 0.09, 0.21) | 0.26 | ( 0.18, 0.35) | 0.19 | ( 0.04, 0.34) | 0.16 | ( 0.09, 0.25) | 0.10 | ( -0.01, 0.22) |
| Total effect | 0.27 | ( -0.22, 0.57) | 0.28 | ( -0.05, 0.60) | 0.32 | ( -0.25, 0.81) | 0.31 | ( -0.13, 0.67) | 0.31 | ( -0.27, 0.85) | 0.31 | ( -0.19, 0.72) |
| Prop. Mediated | 0.60 |  | 0.57 |  | 0.80 |  | 0.61 |  | 0.52 |  | 0.33 |  |
| **Change in ADLs** |  |  |  |  |  |  |  |  |  |  |  |  |
| Indirect effect | 0.07 | ( 0.04, 0.09) | 0.03 | ( 0.00, 0.06) | 0.10 | ( 0.05, 0.15) | 0.09 | ( 0.01, 0.20) | 0.08 | ( 0.04, 0.14) | 0.08 | ( -0.01, 0.19) |
| Total effect | 0.39 | ( 0.18, 0.56) | 0.41 | ( 0.19, 0.62) | 0.10 | ( -0.23, 0.45) | 0.10 | ( -0.23, 0.39) | 0.11 | ( -0.25, 0.35) | 0.11 | ( -0.26, 0.40) |
| Prop. Mediated | 0.17 |  | 0.07 |  | 0.92 |  | 0.86 |  | 0.75 |  | 0.73 |  |
| **Change in Chronic Conditions** |  |  |  |  |  |  |  |  |  |  |  |  |
| Total Effect | -0.07 | ( -0.27, 0.07) | -0.07 | ( -0.23, 0.10) | -0.19 | ( -0.44, 0.12) | -0.19 | ( -0.51, 0.09) | -0.18 | ( -0.46, 0.11) | -0.19 | ( -0.46, 0.10) |
| **Change in Self-Rated Health** |  |  |  |  |  |  |  |  |  |  |  |  |
| Indirect effect | 0.02 | ( 0.01, 0.03) | 0.01 | ( 0.00, 0.02) | 0.02 | ( 0.01, 0.03) | 0.01 | ( -0.01, 0.03) | 0.02 | ( 0.01, 0.03) | 0.02 | ( 0.00, 0.03) |
| Total effect | 0.10 | ( 0.06, 0.15) | 0.11 | ( 0.06, 0.15) | 0.10 | ( 0.03, 0.16) | 0.09 | ( 0.03, 0.16) | 0.10 | ( 0.03, 0.17) | 0.10 | ( 0.03, 0.16) |
| Prop. Mediated | 0.18 |  | 0.08 |  | 0.22 |  | 0.08 |  | 0.22 |  | 0.17 |  |

### II. DNA-methylation analysis

We analyzed DNA-methylation measures of aging in n=3928 Venous Blood Study participants who were included in the DNA-methylation subsample (VBS-DNAm; 42% male, 75% White, 17% Black, aged 50-90 years, mean age=70, SD=9). Participants’ DNA-methylation clock ages were highly correlated with their chronological ages (r>0.72). Participants’ DunedinPoAm values indicated they were aging 7% faster than the rate expected for young-midlife adults (i.e. 1 year of biological change per chronological year; M=1.07, SD=0.09).

#### Comparison of DNA-methylation and blood-chemistry measures of aging

We compared DNA-methylation and blood-chemistry measures of aging. DNA-methylation measures of aging (clock age residuals and DunedinPoAm) were weakly to moderately correlated with one another (r<=0.64). Correlations of DNA-methylation measures of aging with blood-chemistry measures were varied. Horvath-clock age-residuals were not correlated with blood-chemistry measures of aging (r<0.1); Hannum-clock age-residuals were weakly correlated with blood chemistry measures (r=0.1-0.2); correlations were somewhat stronger for PhenoAge-clock residuals (r=0.2-0.3), GrimAge-clock residuals (r=0.3-0.4), and DunedinPoAm (r=0.2-0.3). Correlations and scatterplots are shown in **Supplemental Figure S1**.

#### Older adults with more advanced/faster biological aging in DNA-methylation analysis showed deficits in healthspan-related characteristics and were at increased risk of mortality

Participants with more advanced/faster biological aging measured from DNA-methylation had poorer outcomes for all healthspan-related characteristics and increased risk for mortality. Of the clocks, which measure how much a person has aged up to the time of measurement, effect-sizes were largest for GrimAge (IRR=1.20-1.39 per SD for functional impairments and prevalent and incident ADLs and chronic conditions; r=0.32 for self-rated health and r=0.08 for change in self-rated health; HR=2.32 for mortality per SD). Effect-sizes were somewhat smaller for the PhenoAge and Hannum clocks. For the Horvath clock, effect-sizes were smaller and often not statistically different from zero at the alpha=0.05 level. For DunedinPoAm pace of aging, which measures how rapidly a person is aging at the time of measurement, effect-sizes were smaller than for GrimAge and somewhat larger than for the other clocks (IRR=1.14-1.27 per SD for functional impairments and prevalent and incident ADLs and chronic conditions, r=0.22 for self-rated health and r=0.06 for change in self-rated health, and HR=1.71 for mortality per SD). Effect-sizes are reported in **Figure 1** and **Supplemental Table S2**.

#### Black HRS VBS-DNAm participants showed deficits in healthspan-related characteristics and increased risk for mortality as compared with White participants

Black participants more often demonstrated functional impairments (IRR=1.19 [1.05-1.35]) and reported more ADLs (IRR=1.81 [1.57-2.07]) and diagnosed chronic conditions (IRR=1.24 [1.16-1.33]) and poorer self-rated health (d=0.28 [0.19-0.37]) as compared with White participants. Over follow-up from 2016 baseline, Black participants had higher mortality (HR=1.34 [0.82-2.17]) and were more likely to report incident ADLs (IRR=1.11 [0.90-1.37]) and declines in self-rated health (d=0.10 [0.03-0.17]), although, with the exception of changes in self-rated health, associations were not statistically different from zero at the alpha=0.05 level. As in the full VBS sample and Black-White differences were in the opposite direction for incident chronic conditions (IRR=0.84 [0.64-1.10]). Effect-sizes are reported in **Supplemental Table S3**.

#### Black HRS VBS-DNAm participants showed more advanced biological aging according to the GrimAge clock and faster biological aging according to DunedinPoAm pace of aging as compared with White participants

In analysis of Black-White disparities, the GrimAge clock and the DunedinPoAm pace of aging indicated more advanced/faster biological aging in Black as compared with White participants (GrimAge d=0.36, 95% CI=[0.25,0.48]; DunedinPoAm d=0.38, 95% CI=[0.24,0.51]). In contrast, the 1^st^ generation Horvath and Hannum clocks and the PhenoAge Clock did not (Horvath-clock d=-0.02, 95% CI=[-0.10,0.07]; Hannum-clock d=-0.37, 95% CI=[-0.49,-0.26]; PhenoAge clock d=0.08, 95% CI=[-0.04,0.20]). Effect-sizes are reported in (**Supplemental Table S1, Figure 2**).

#### The GrimAge clock and DunedinPoAm pace of aging DNA-methylation quantifications of biological aging accounted for significant proportions of Black-White differences in healthspan-related characteristics and mortality risk

The Horvath, Hannum, and PhenoAge clocks did not indicate more advanced aging in Black as compared to White participants and were not included in mediation analysis. More advanced GrimAge and faster DunedinPoAm pace of aging in Black as compared with White participants mediated 13-43% of Black-White differences in functional impairment, ADLs, chronic conditions, and self-rated health in cross-sectional analysis and 22-92% of Black-White differences in ADL incidence, change in self-rated health, and mortality risk in longitudinal analysis. Effect-size estimates for Black-White differences in healthspan-related characteristics before and after adjustment for GrimAge clock and DunedinPoAm pace of aging measures are reported in **Supplemental Table S3**. Mediation analysis results are reported in **Table 3** and **Supplemental Table S4**.

### III. Sensitivity Analysis

Standard mediation analysis assumes that the association of the mediator with the outcome is consistent across levels of exposure. We conducted sensitivity analysis to evaluate this assumption and to test the robustness of mediation results when this assumption was relaxed. Overall, association magnitudes tended to be smaller for analysis of Black as compared to White participants (**Supplemental Tables S2 and S5**). When we relaxed the mediation-analysis assumption that association magnitudes were the same for Black and White subsamples, mediation proportions were reduced. The magnitude of this reduction varied from near zero to as much as 50% (“E-M interaction” columns of **Table 3** and **Supplemental Table S4**). The sensitivity analysis is described in detail in **Supplemental Methods Section 3**.

## DISCUSSION

We investigated biological aging as a mediator of Black-White disparities in healthspan-related characteristics in the US Health and Retirement Study. In the HRS VBS and VBS-DNAm samples, Black participants experienced more functional impairments, more difficulties with activities of daily living (ADLs) and higher incidence of ADLs over follow-up, more chronic-disease diagnoses, poorer self-rated health and greater declines in self-rated health over follow-up, and increased risk for mortality as compared to White participants. Black participants also showed more advanced/faster biological aging based on the three blood-chemistry measures we analyzed, the DNA-methylation GrimAge clock, and the DunedinPoAm pace of aging measure. In mediation analysis, these measures of more advanced/faster biological aging accounted for up to 95% of Black-White differences in healthspan-related characteristics and mortality. These findings are consistent with the weathering hypothesis that racially patterned determinants of health accelerate biological aging, contributing to Black-White disparities in healthspan (19, 20), and provide limited proof-of-concept for use of quantifications of biological aging in health disparities research.

Measures of biological aging have been suggested as surrogate endpoints for trials testing therapies to prolong healthy lifespan based on evidence that they predict aging-related changes in health, functioning, and mortality risk (36-38). No measures of biological aging have yet been tested in randomized clinical trials that include measurements of primary endpoints related to healthy lifespan. As a result, all fall short of the US Food and Drug Administration’s criterion that validated surrogate endpoints be reliable predictors of clinical benefit (39).

Our findings offer mixed support for the measures we studied as candidate surrogate endpoints (i.e. proposed surrogates for which prediction of clinical benefit is not yet established) (39). Specifically, a criterion for a candidate surrogate endpoint is the robustness of associations between the candidate surrogate and primary outcomes across population subgroups. This robustness is not yet established for measures of biological aging, especially in the case of DNA-methylation-based measures, for which algorithms were developed using data from mostly White-European samples, e.g. (40). Our analysis found mixed support for a hypothesis of consistent associations across Black and White older adults. Effect-sizes tended to be smaller for analyses of Black as compared with White participants. However, the same Black-White differences in effect-sizes were also observed for analysis of chronological age, indicating that differential precision of aging measures between Black and White participants was likely not the cause. Instead, our data may reflect that Black Americans are disproportionately subject to non-aging causes of disease and disability such as injury or accidents, which would not be captured in measures of biological aging. Therefore, while our findings do not yet establish validity of biological aging measures as candidate surrogate endpoints, they do support cautious interpretation of group differences in these measures of biological aging as evidence of disparities in healthy aging.

Our findings suggest guidance to future studies. The conceptual model guiding our study proposes that Black-White disparities in healthspan arise from faster/more-advanced biological aging in Black as compared to White Americans. In our analysis, biological aging measurements from the Horvath, Hannum, and PhenoAge clocks did not fit this model; they showed no Black-White differences or differences in the opposite direction. These clocks may not be well-suited to characterizing Black-White disparities in healthy aging. The HRS result that PhenoAge clock values did not differ between Black and White participants contrasts with a report from the Women’s Health Initiative showing more advanced biological aging in Black as compared with White women using this measure (23). Follow-up in additional cohorts is needed. Results also contribute evidence that first-generation DNA-methylation clocks—developed to predict chronological age—are both less predictive of healthspan-related characteristics and less sensitive to exposures that shorten healthy lifespan, as compared to blood-chemistry-derived measures, newer DNA-methylation clocks developed to predict mortality, and the DunedinPoAm DNA-methylation pace of aging (26, 28, 41-43). Future studies using DNA-methylation to investigate biological aging as a mediator between risk exposures and healthy-aging phenotypes, especially in the context of health disparities, may be best served by focus on 2^nd^-generation DNA methylation clocks, especially the GrimAge clock, and DunedinPoAm pace of aging measures.

We acknowledge limitations. There is no gold standard measure of the construct of biological aging (44). Our conclusions regarding biological aging as a candidate mediator of health disparities could be specific to the measures we analyzed. However, consistent evidence across different biological substrates and measurement methods builds confidence that results do reflect aging processes. DNA-methylation measures of aging may reflect variation in the white blood cell composition of samples from which DNA is extracted (45). Sensitivity of our results to this variation could not be tested because the HRS has not yet released whole-genome DNA-methylation data or estimates of white blood cell proportions. Mortality selection may bias results toward the null. Many individuals born in the same years as the HRS participants whose data we analyzed will not have survived to the time of HRS data collection, especially Black Americans, who face shorter life expectancies as compare to White Americans (46). If survivors aged more slowly than those who died at younger ages, our analysis could underestimate Black-White differences in biological aging and healthspan. Our estimates of disparities are therefore likely to be conservative. Indirect-effect estimates in our mediation models are conditional on the assumption that there are no common causes of biological aging and healthspan-related characteristics omitted from the model. To the extent that these causes exist, our estimates of the proportion mediated may be biased upwards. Finally, there is the possibility of detection and reporting bias. For example, racial disparities in chronic disease might be underestimated if White participants were more likely to be diagnosed due to greater access to health care (47, 48).

Within the bounds of these limitations, our findings have implications for future research and public health surveillance. More advanced/faster biological aging in Black as compared to White HRS participants and the potential role of these differences in mediating Black-White health disparities highlight the need for studies of when and how Black-White differences in biological aging arise. Life-course longitudinal studies are needed to establish when aging trajectories begin to diverge for Black and White Americans. Studies are also needed to identify life-course phenomena through which racism and socioeconomic resource differentials drive faster aging in Black as compared to White Americans; maternal and perinatal health, social exclusion and victimization of young adults, occupational exposures to young and midlife adults, and access to healthcare later in life all represent potential drivers of Black-White disparities in aging.

Our results suggest promise for the application of biological-aging measures for evaluating and monitoring Black-White disparities in healthy aging, in particular the GrimAge clock and DunedinPoAm pace of aging, with the caveat that these measures do not provide a complete summary of processes driving Black-White health disparities. These same measures can, in parallel, provide new outcome measures for evaluations of social policy experiments. A primary application of biological aging measures within the emerging field of geroscience is to provide surrogate endpoints for extension of healthy lifespan (36, 49). Results from this study suggest they may also have utility in trials of interventions that aim to eliminate health disparities by repairing inequalities in social determinants of health.

## Data Availability

Data is available from the University of Michigan (https://hrs.isr.umich.edu/data-products)

## Acknowledgements

This research was supported by National Institute on Aging Grants R01AG061378 and R01AG066887, Russell Sage Foundation Grant 1810-08987, and the Jacobs Foundation. CLC is supported by a fellowship from the National Institute of Mental Health (5T32MH013043). DWB is a fellow of the CIFAR CBD Network.

