## Supplemental Methods and Figures for "Testing Black-White disparities in biological aging in older adults in the United States: analysis of DNA-methylation and blood-chemistry methods"

**1. Measurements of Biological Aging**

**Blood-Chemistry Methods**

In blood-chemistry analysis, we focused on three algorithms developed for blood chemistry analytes routinely measured in clinical settings and which have been most commonly used in studies to date: the recently published "Phenotypic Age" measure, Klemera-Doubal Method Biological Age, and Homeostatic Dysregulation.

The PhenoAge algorithm was developed from elastic-net Gompertz-regression of mortality on 42 blood-chemistry markers and chronological age in the NHANES III dataset (1). The resulting algorithm comprised 9 biomarkers (albumin, alkaline phosphatase, creatinine, C-reactive protein (log), glucose, white blood cell count, lymphocyte %, mean cell volume, and red cell distribution width) and chronological age. The elastic-net derived algorithm computes a predicted hazard of mortality, referred to as a mortality score. This mortality score is converted to a biological age based on comparison to predicted hazards from a univariate Gompertz regression of mortality on chronological age. Thus, a participant’s Phenotypic Age is the chronological age at which the participant’s mortality hazard would match the norm in the original NHANES III sample.

Klemera-Doubal method (KDM) Biological Age measures aging from the association of physiological parameters with chronological age (2). Whereas Phenotypic Age is the age at which a person’s predicted mortality risk is at the norm in the reference population, KDM Biological Age is the age at which a person’s physiology is at the norm in the reference population. For our analysis, we defined the reference population as NHANES III participants aged 30-75 (3, 4).

Homeostatic dysregulation measures aging as how deviant a person’s physiology is from a reference norm as computed by Mahalanobis distance (5). The homeostatic dysregulation method does not estimate an age value. Instead, the Mahalanobis distance is the estimate of aging. We defined the homeostatic dysregulation reference norm from NHANES III participants aged 20-30, non-obese, and for whom all biomarkers were within the clinically normal range (3, 6).

**DNA-Methylation Methods**

In DNA-methylation analysis, we considered five different measures computed by the HRS (7). Four of these are known as epigenetic clocks, DNA-methylation algorithms that produce values that are interpretable as ages. Clock-ages that are older than the chronological age of the person being measured indicate an advanced state of biological aging; clock-ages younger than chronological age indicate delayed aging. We analyzed epigenetic clocks proposed by Horvath, Hannum et al., Levine et al., and Lu et al. The Horvath and Hannum et al. clocks were developed using machine learning analysis to predict chronological age (8, 9). These clocks are referred to as “first-generation epigenetic clocks.” The clock proposed by Levine et al., known as the PhenoAge Clock, and the clock proposed by Lu et al., known as the GrimAge clock, were developed to predict aging-related mortality (1, 10). These clocks are referred to as “second-generation epigenetic clocks.” The PhenoAge clock is a DNA-methylation prediction of the blood-chemistry PhenoAge described above. The GrimAge clock was developed by first defining DNA-methylation algorithms to predict several blood proteins, smoking, and chronological age, and then fitting these DNA-methylation biomarkers to mortality risk. For analysis, participants’ clock-ages were regressed on their chronological ages and residual values were calculated. We refer to these measures hereafter as “clock-age residuals.”

The fifth DNA-methylation algorithm measures pace of aging. Pace of aging is the rate of decline in system integrity that occurs with advancing chronological age. The measure we analyzed, DunedinPoAm, was developed from analysis of longitudinal change in 18 biomarkers tracking multi-organ-system integrity in a birth cohort followed from age 26-38 years (11). Values are interpretable as years of physiological change occurring per 12-month calendar interval in healthy adults. Dunedin PoAm values above one indicate a faster than normal pace of aging; values below one indicate a slower pace of aging.

**2. Mediation Analysis.**

We tested mediation of Black-White disparities in biological aging using a regression-based approach as described by Valeri and VanderWeele (12). This method allows for the decomposition of the total effect of the exposure on the outcome of interest through direct and indirect pathways. We specified two regressions for each relationship of interest, $E\left( Y \right|t, m, c)$and $E(M|t,c)$. In these expectation statements, $Y$ is the outcome, here a healthspan-related characteristic, $M$ is the mediator, here biological-age advancement, and $t$ is the exposure of interest, here a social determinant of health. We modeled $E\left( Y \right|t, m)$using Poisson regression to estimate IRRs for the healthspan-related characteristics, which were operationalized as counts. We modeled $E(M|t)$ using linear regression of the continuously-distributed measure of biological age advancement on the social determinant of health (Black versus White racial identity). Models included sex, chronological age, and region of residence as covariates. Indirect effect estimates in mediation models are conditional on the assumption that there are no common causes of biological aging and healthspan-related characteristics omitted from the model. To the extent that these causes exist, our estimates of the proportion mediation may be biased upwards. We used the R package *CMAverse* (13) to estimate direct and indirect effects based on these regressions. We calculated confidence intervals using bootstrapping to obtain standard error estimates. We tested robustness of mediation results to potential exposure-mediator interactions following the approaches outlined by Valeri and VanderWeele (12, 14).

**3. Sensitivity Analysis of Exposure-Mediator Interactions**

Mediation analysis assumes that the association of the mediator with the outcome is consistent across levels of exposure. We conducted sensitivity analysis to evaluate this assumption and to test the robustness of mediation results when this assumption was relaxed. First, we fitted regressions of healthspan characteristics and mortality on measures of biological aging in the Black and White subsamples of HRS VBS and VBS-DNAm samples. Next, we conducted tests of super-multiplicative and super-additive interaction between the exposure (Black vs. White racial identity) and the mediators (measures of biological aging) in models predicting healthspan characteristics and mortality. Finally, we re-estimated mediation models allowing for exposure-mediator interactions using the model proposed by Valeri and colleagues (14).

Effect-sizes tended to be smaller for analysis of Black as compared with White HRS participants. Stratified regression results are reported in **Supplemental Table 2**.

Tests of interaction were inconsistent. We formally tested exposure-mediator interactions between race and biological-aging by fitting models including a product term testing the interaction between race/ethnicity and biological aging. We conducted analysis of the three blood-chemistry measures and two DNA-methylation measures we tested in mediation analysis. We tested for heterogeneity by evaluating interactions between race and biological aging on multiplicative and additive scales. Multiplicative interaction was tested from the coefficient estimate for the product term the regression models. Additive interaction was tested using the Relative Excess Risk due to Interaction (RERI) method. In this method, the total combined effect of race and biological-age advancement $e^{\beta_{1}+\beta_{2}+\beta_{3}}$ is compared with the sum of the individual effects of race and biological-age advancement $e^{\beta_{1}}+e^{\beta_{2}}$. This comparison is made by assessing $RERI=(e^{\beta_{1}+\beta_{2}+\beta_{3}})-(e^{\beta_{1}} )-(e^{\beta_{2}})+1$. If the RERI is equal to zero, no additive interaction is present. If the RERI is greater or less than zero, super-additivity and subadditivity are implied. For analysis of functional impairments, prevalent and incident ADLs and chronic conditions, and mortality, tests of super-additive interaction (RERI analysis) mostly failed to reject the null hypothesis that associations were similar in Black and White participants at the alpha=0.05 level. In contrast, tests of super-multiplicative interaction (product-term coefficient analysis) rejected the null hypothesis at the alpha=0.05 level for most analysis of prevalent ADLs and chronic conditions and for incident ADLs in the case of the blood chemistry measures of biological aging. For analysis of self-rated health, super-additive interaction was tested by the product-term in the regression models. This test rejected the null hypothesis of no difference between Black and White subsamples in analysis of blood chemistry measures, but not DNA-methylation measures. Tests of interaction are reported in **Supplemental Table S5**.

One possible cause of differences in association magnitudes between Black and White participants is that biological aging measures were developed in samples that were majority or entirely White (**Table 1**). A consequence may be that measurements are less precise in Black as compared with White Americans. However, standard deviations of biological aging measures were similar for Black and White participants (**Supplemental Table S1**). To further test this possibility, we repeated our interaction analysis using chronological age, which should be measured with identical precision in Black and White participants, instead of measures of biological aging. The pattern of results for chronological age was similar to the results for the DNA-methylation measures; tests of super-additive interaction failed to reject the null-hypothesis that associations were similar in Black and White subsamples; tests of super-multiplicative interaction rejected the null at the alpha=0.05 level for analysis of prevalent ADLs and chronic conditions (**Supplemental Table S5**). In addition, the null hypothesis was also rejected for analysis of prevalent functional impairments. Thus, differential measurement precision is likely not the cause of Black-White differences in magnitudes of association between biological aging measures and healthspan characteristics.

An alternative explanation for the relatively smaller association magnitudes in Black as compared with White participants is that non-aging causes of healthspan-related characteristics, such as injuries and accidents, are more common in Black as compared to White participants. Such non-aging causes would drive poorer healthspan-related characteristics in the Black subsample, regardless of their state and pace of aging, in turn constraining variation in outcomes reducing association magnitudes (1). **Supplemental Figure S2**, which plots predicted values of healthspan-related characteristics across distributions of biological aging measures, presents some evidence consistent with this explanation: The plots show that the principal difference between risk gradients for Black and White participants is that Black participants are at a higher level of risk even when their biological aging is relatively less-advanced/slower than the norm.

Such a phenomenon can be statistically represented within our mediation models as exposure-mediator interaction. We tested sensitivity of our results to potential exposure-mediator interactions using the model proposed by Valeri and colleagues (14). Results are reported in the “With E-M Interaction” columns of **Table 3** and **Supplemental Table S4**.

**Supplemental Methods Figure 1. Directed Acyclic Graph (DAG) depicting potential causes of smaller effect-sizes for associations of biological aging measures with healthspan characteristics in Black as compared to White participants.** The figure is based on the DAG in figure 2C in le Cessie (2012, Epidemiology). The node labeled “Biological Aging” represents the unobserved process of biological aging. The node labeled “Biological Aging*” represents the measured variable in analysis. The measured variable is caused by the unobserved process of aging and measurement error, including systematic measurement error patterned by the exposure (path A) and non-systematic measurement error (represented by U). The DAG identifies two potential causes of differences in effect-sizes between Black and White participants. Path A shows measurement error in the mediator conditional on exposure status (the edge connecting social determinants of health and measured biological aging). Such error could arise, for example, because samples used to develop biological aging measures included few Black participants. Path B shows non-aging causes of healthspan, such as injuries or accidents, that may be patterned across levels of exposure.

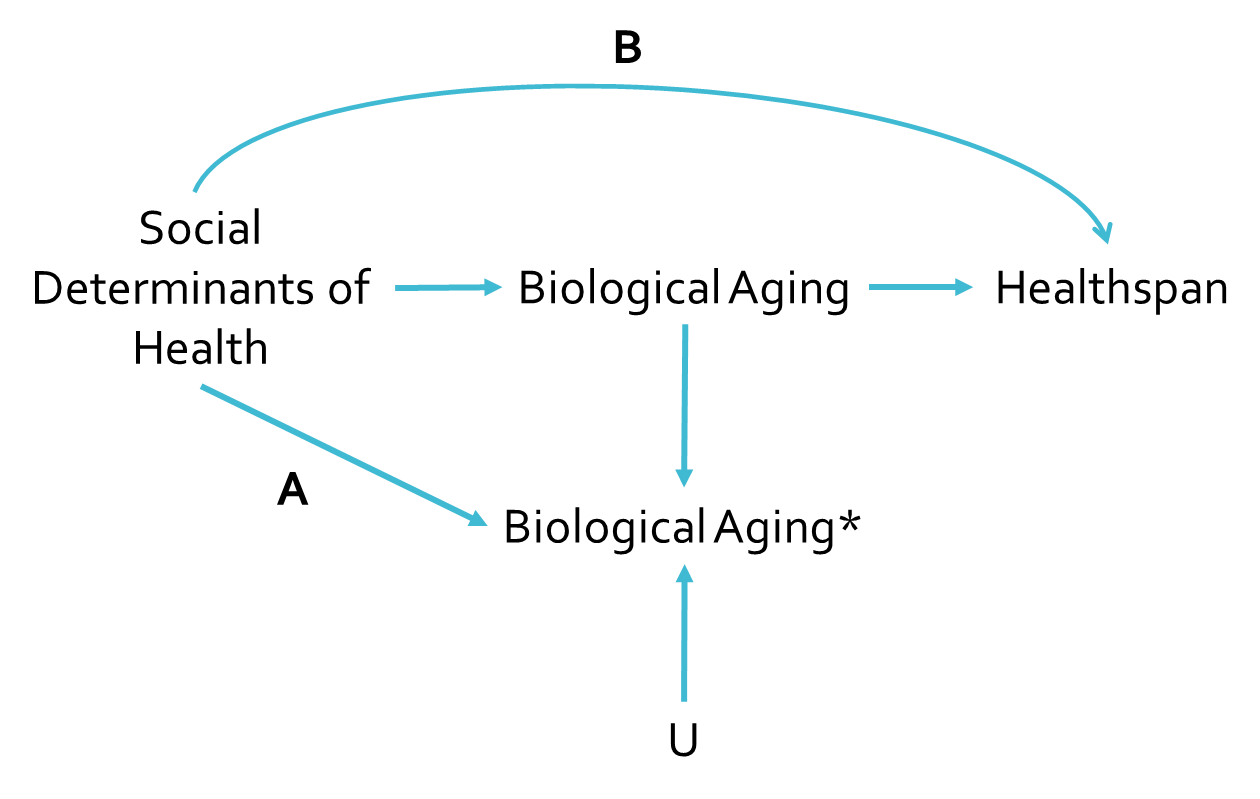

**Supplemental Methods Table 1. Measurement of healthspan-related characteristics.** This table describes the construction of each healthspan-related characteristic used in cross-sectional analyses of Black and White participants in the 2016 HRS VBS and VBS-DNAm subsamples.

| **Panel A. Cross-sectional measures of prevalent healthspan-related characteristics** | |
| --- | --- |
| **Functional Impairments** (VBS n=3621, VBS-DNAm n=1596) | Functional limitations were measured from tests of lung function (Peak Flow), grip strength, gait speed, and balance. The tests are described in detail below. We classified those with test scores in at the 30^th^ percentile of the HRS distribution or lower as having poor performance. Percentiles for grip strength were calculated separately for men and women. We computed functional impairment scores for participants providing data on at least 3 of the 4 tests. We summed the number of tests on which participants demonstrated poor performance and coded the resulting functional impairment score as 0, 1, 2, or 3+ functions with limitation. |
| Peak Flow (L/min) | Participants in the physical exam were administered the breathing test using a Mini-Wright peak flow meter with a disposable mouthpiece (Clement Clarke International Ltd., Harlow, United Kingdom). Three measurements were conducted, each 30 seconds apart. The mean of the three peak flow tests was used as a summary measure of peak flow. |
| Grip Strength (kg) | Participants 65+ years of age in the physical exam were administered the grip strength test. Grip strength was measured with a Smedley spring-type hand dynamometer (Stoelting Company, Wood Dale, Illinois), and results were recorded each time to the nearest 0.5 kg. The maximum grip strength recorded across 2 trials was used as a summary measure of grip strength. |
| Walk Speed (log) | Participants 65+ years of age in the physical exam were administered the walking speed test. The test was conducted in a non-carpeted area on a straight walking path 98.5 inches long. The walking speed test was conducted twice- once in each direction. Time to test completion was recorded in seconds, to two decimal places, using a stopwatch. Participants were instructed to use a walking stick or other aid if necessary. The minimum time to complete the walking speed test was used as a summary measure of walk speed. This summary measure was then reverse coded and log-transformed for analysis. Higher scores reflect a faster walking speed |
| Balance | All participants in the physical exam were asked to perform a series of balance tests. Participants were first administered the semi-tandem balance test. Those who did not pass the semi-tandem test were administered the side-by-side test; those who passed were administered the full tandem balance test. In the semi-tandem test, participants were asked to stand with heel of one foot touching the big toe of the other foot for 10 seconds. In the side-by-side test, participants were asked to stand with their feet together for 10 seconds. In the full tandem test, participants were asked to stand with their feet together for 30 seconds if 65 or older, and 60 if younger than 65. For all tests, participants were told they could use their arms, bend their knees, or move their body to maintain their balance, but not to move their feet.  We assigned participants a balance score based on whether they were able to complete the balance tests administered:  1 = did not complete semi-tandem and did not complete side-by-side  2 = did not complete semi-tandem but completed side-by-side  3 = completed semi-tandem but did not complete full tandem  4 = completed semi-tandem and full tandem |
| **Chronic Conditions** (VBS n=8196, VBS-DNAm n=3597) | Participants were asked if they had ever been diagnosed with each of the following six (6) chronic diseases: hypertension, type II diabetes, cancer (excluding minor skin cancer), chronic lung disease, heart problems (heart attack, coronary heart disease, angina, congestive heart failure), and/or stroke. We summed the number of comorbidities reported by each participant to obtain a total comorbidity score. Comorbidity score was then categorized as None, 1, 2, or 3+. |
| **Self-Rated Health** (VBS n=8191, VBS-DNAm n=3595) | Participants were asked to rate their own health on a scale of one to five (1=excellent, 5=poor). Participant report was conserved for analysis. |
| **Number of ADLs** (VBS n=8198, VBS-DNAm n=3598) | Participants were asked if they had difficulty performing 6 activities of daily living: dressing, eating (such as cutting up your food), bathing and showering, getting in and out of bed, toileting, and walking across a room. We summed the number of activities for which participants reported difficulty and coded the resulting ADL score as 0, 1, 2, or 3+ ADLs. |
| **Panel B. Longitudinal measures of healthspan-related characteristics and mortality (2016-2018)** | |
| **Mortality** (VBS n=8198, VBS-DNAm n=3598) | HRS collects data on date of death for participants who have died between waves of sample collection. We coded mortality as a binary variable (dead/alive). For time-to-event analyses, we calculated time from follow-up from 2016 by subtracting the 2016 interview date from date of death. |
| **Incident ADLs** (VBS n=7230, VBS-DNAm n=3164) | For all participants who provided data on ADL limitations at both the 2016 and 2018 waves of the HRS, we subtracted the total self-reported number of ADL limitations in 2016 from the total self-reported number of ADL limitations in 2018. Negative values were recoded to 0. Values ranged from 0 to 6. |
| **Incident chronic conditions** (VBS n=7236, VBS-DNAm n=3166) | For all participants who provided data on ever-diagnosed chronic conditions at both the 2016 and 2018 waves of the HRS, we subtracted the total number of physician-diagnosed chronic conditions in 2016 from the total number of physician-diagnosed chronic conditions in 2018. Any discrepancies resulting in negative values were recoded to 0. Values ranged from 0 to 3. |
| **Change in self-rated health** (VBS n=7229, VBS-DNAm n=3163) | For all participants who provided data on self-rated health at both the 2016 and 2018 waves of the HRS, we subtracted self-rated health score in 2016 from self-rated health score in 2018. Values ranged from -4 to 4. |

### Supplemental Figures

**Supplemental Figure S1a. Correlations among three blood-chemistry and five DNA-methylation measures of biological-age advancement among Black and White participants in the US Health and Retirement Study.** Biological aging measure labels are listed on the matrix diagonal. Pearson correlations are shown above the diagonal. Correlations are reported for the biological aging measures listed below and to the left of the cell. Scatterplots and linear fits illustrating associations are shown below the diagonal. The Y axis of the plots corresponds to the biological aging measure to the right of the cell. The X axis of the plots corresponds to the biological aging measure above the cell. Biological-age advancement values were calculated by fitting regressions of biological age measures on chronological age and computing residual values.

**Panel A. Full Sample (Blood-chemistry measures n=9005, DNAm measures n=3928)**

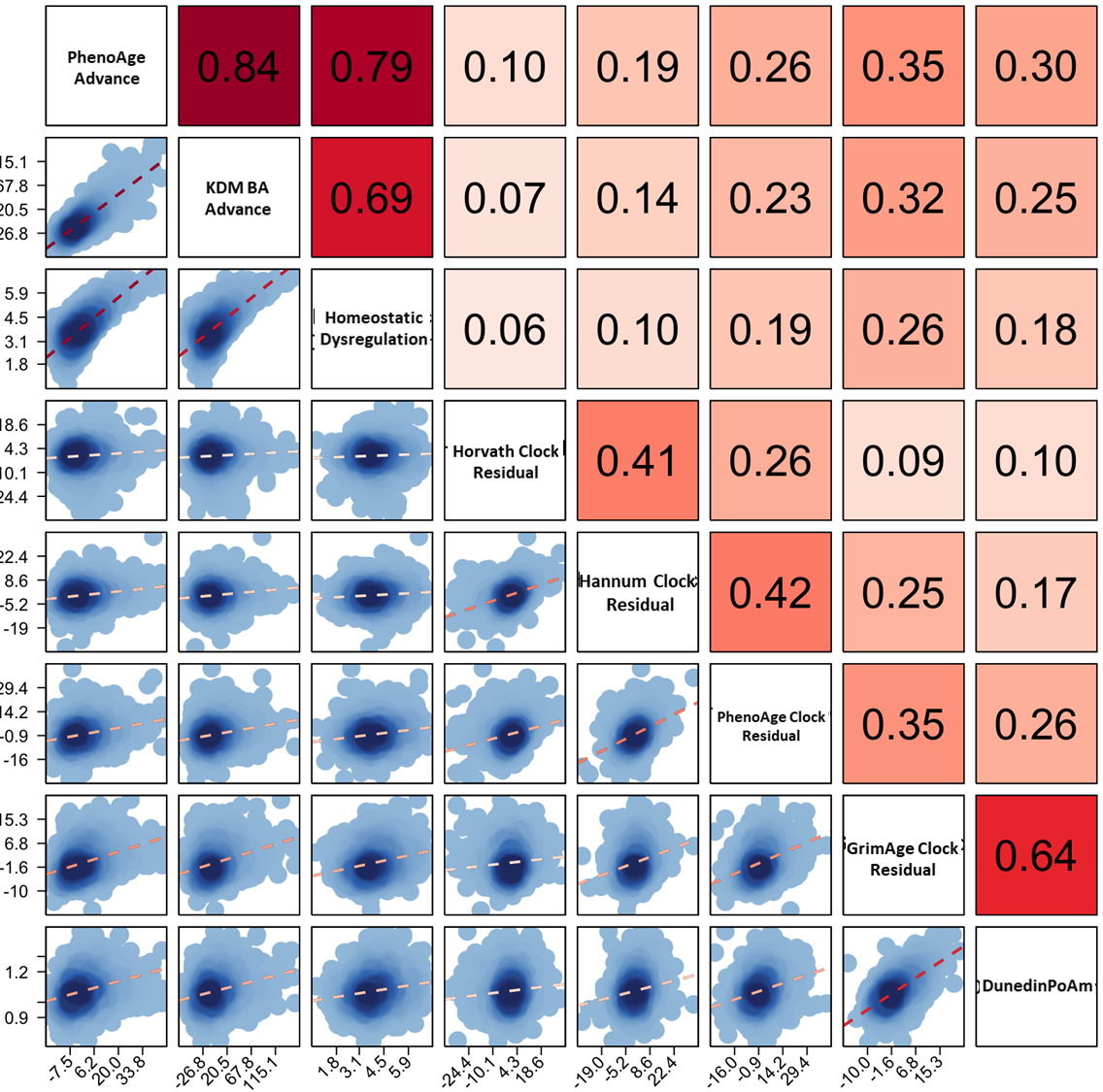

**Panel B. White Participants (Blood-chemistry measures n=6608, DNAm measures n=2930)**

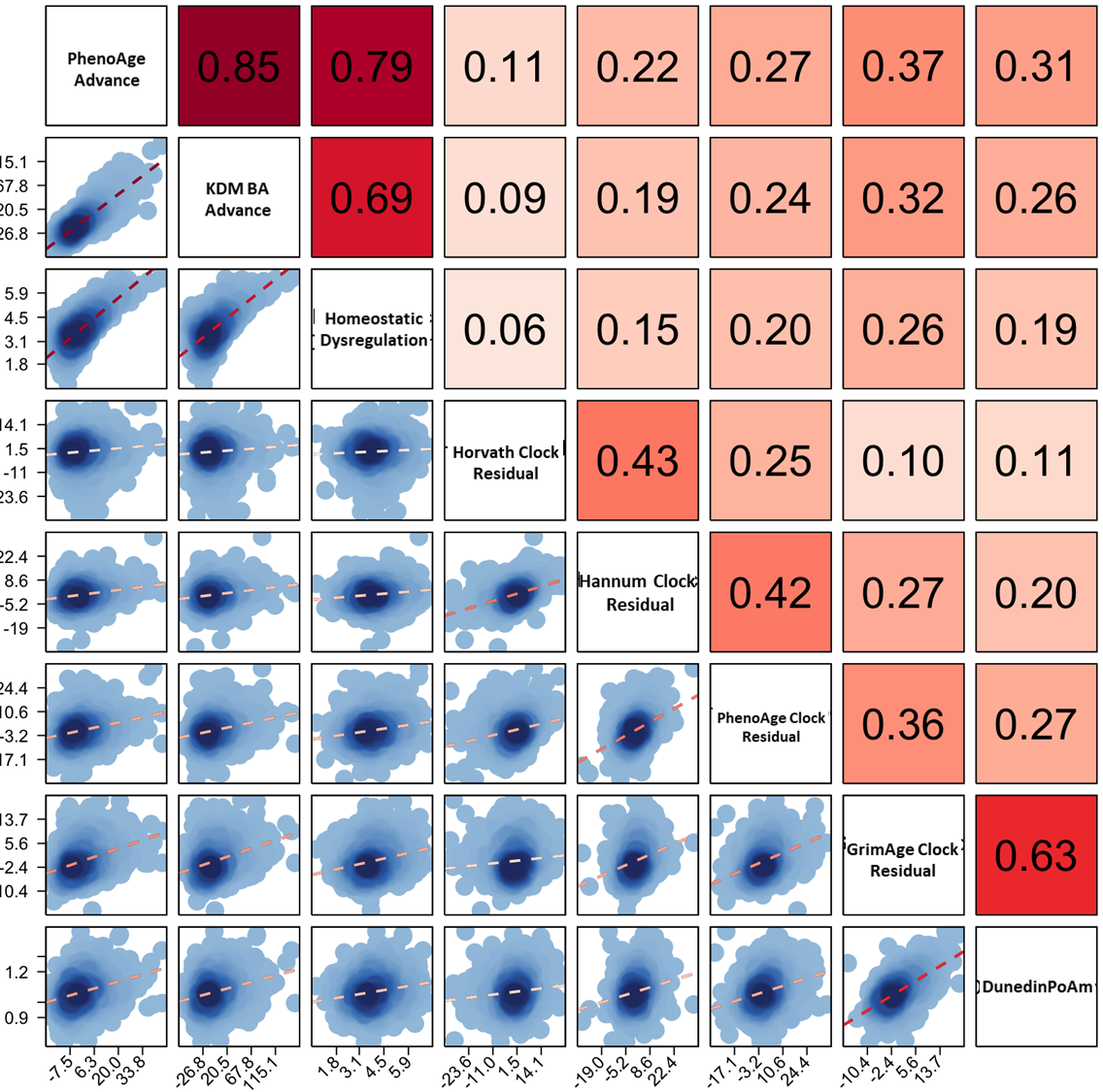

**Panel C. Black Participants (Blood-chemistry measures n=1590, DNAm measures n=668)**

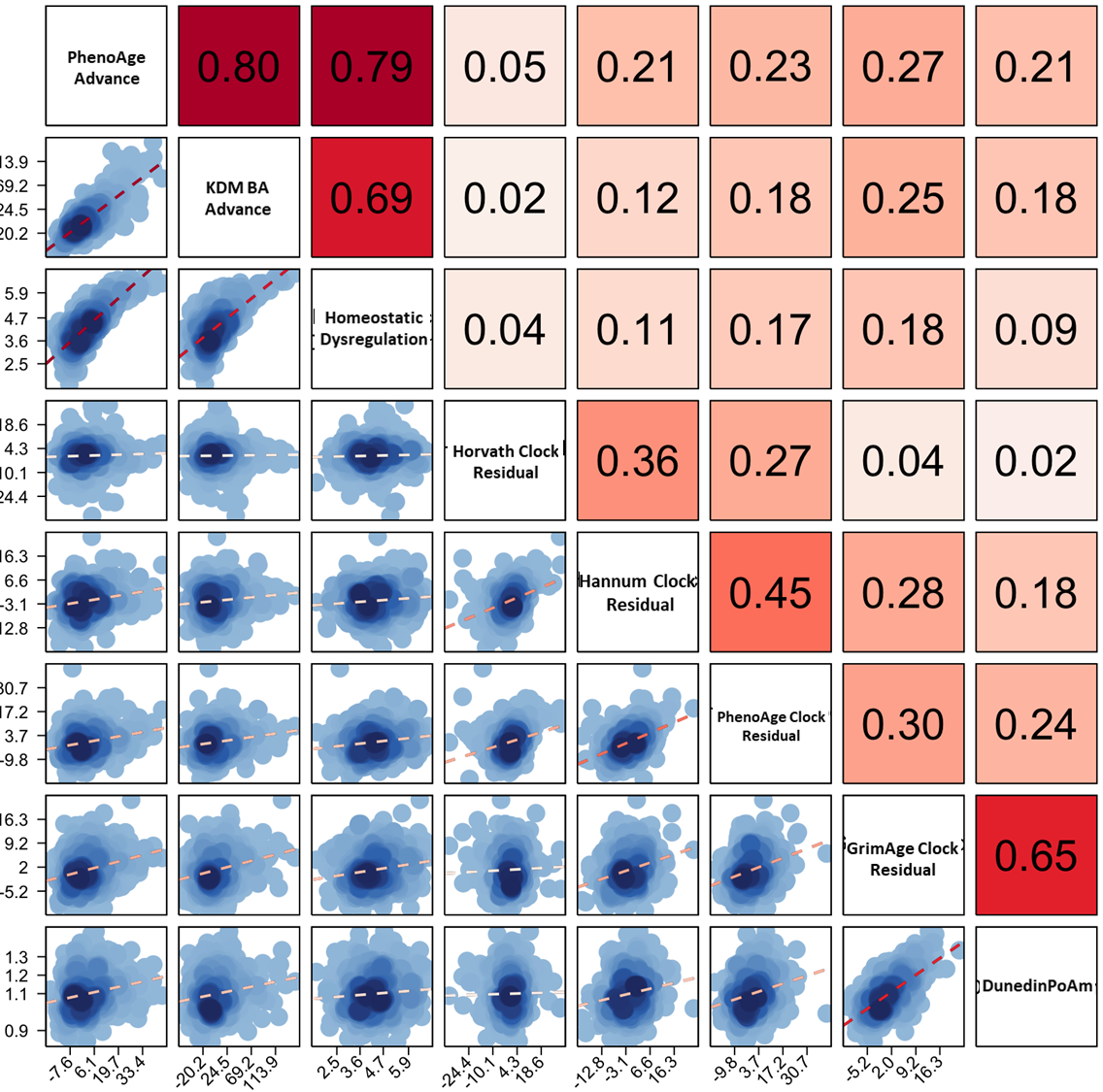

**Supplemental Figure S1b. Correlations among chronological age, three blood-chemistry, and five DNA-methylation measures of biological age among Black and White participants in the US Health and Retirement Study.** Biological age measure labels are listed on the matrix diagonal. Pearson correlations are shown above the diagonal. Correlations are reported for the biological age measures listed below and to the left of the cell. Scatterplots and linear fits illustrating associations are shown below the diagonal. The Y axis of the plots corresponds to the biological age measure to the right of the cell. The X axis of the plots corresponds to the biological age measure above the cell. Biological-age advancement values were calculated by fitting regressions of biological age measures on chronological age and computing residual values.

**Panel A. Full Sample (Blood-chemistry measures n=9005, DNAm measures n=3928)**

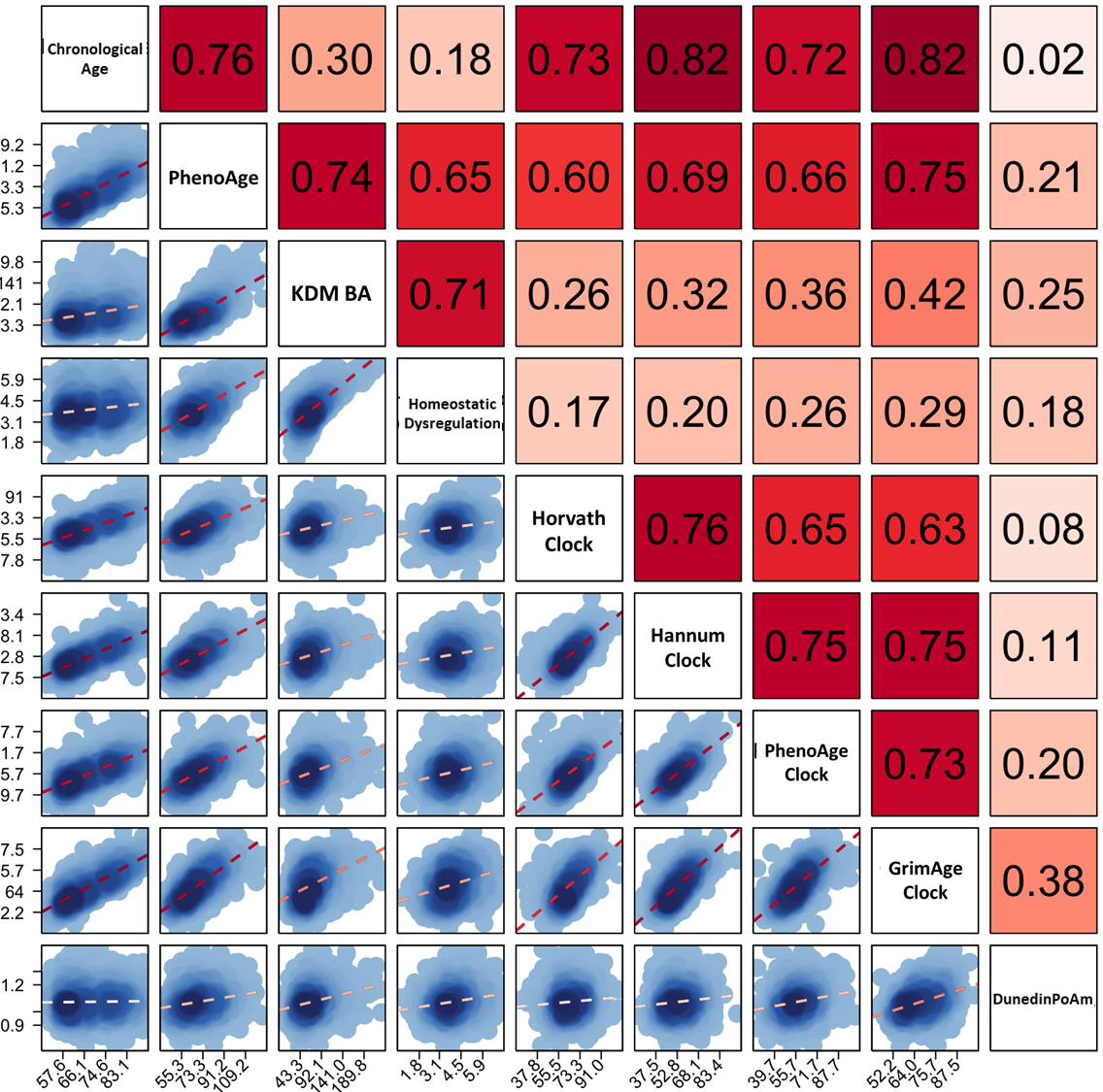

**Panel B. White Participants (Blood-chemistry measures n=6608, DNAm measures n=2930)**

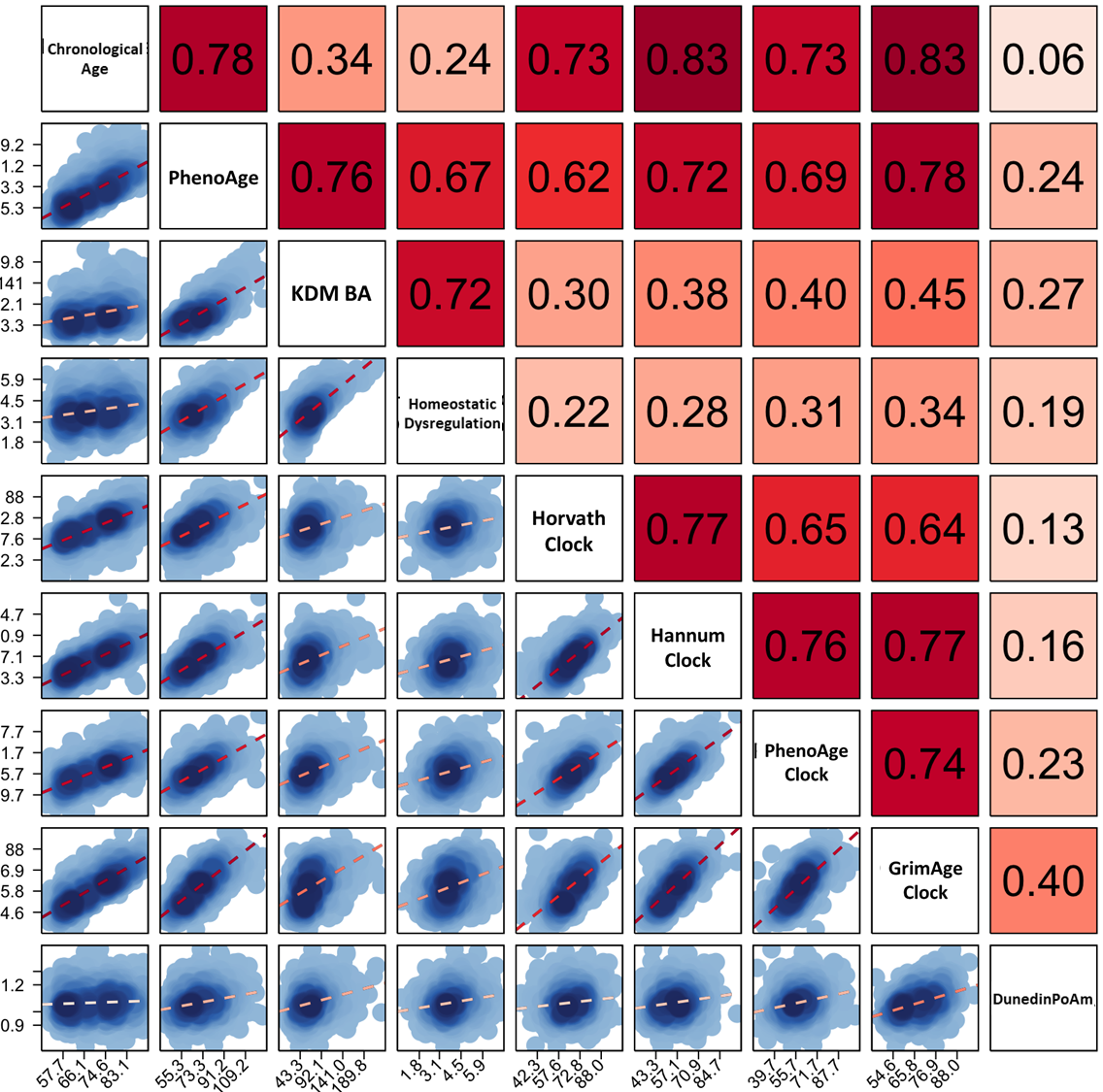

**Panel C. Black Participants (Blood-chemistry measures n=1590, DNAm measures n=668)**

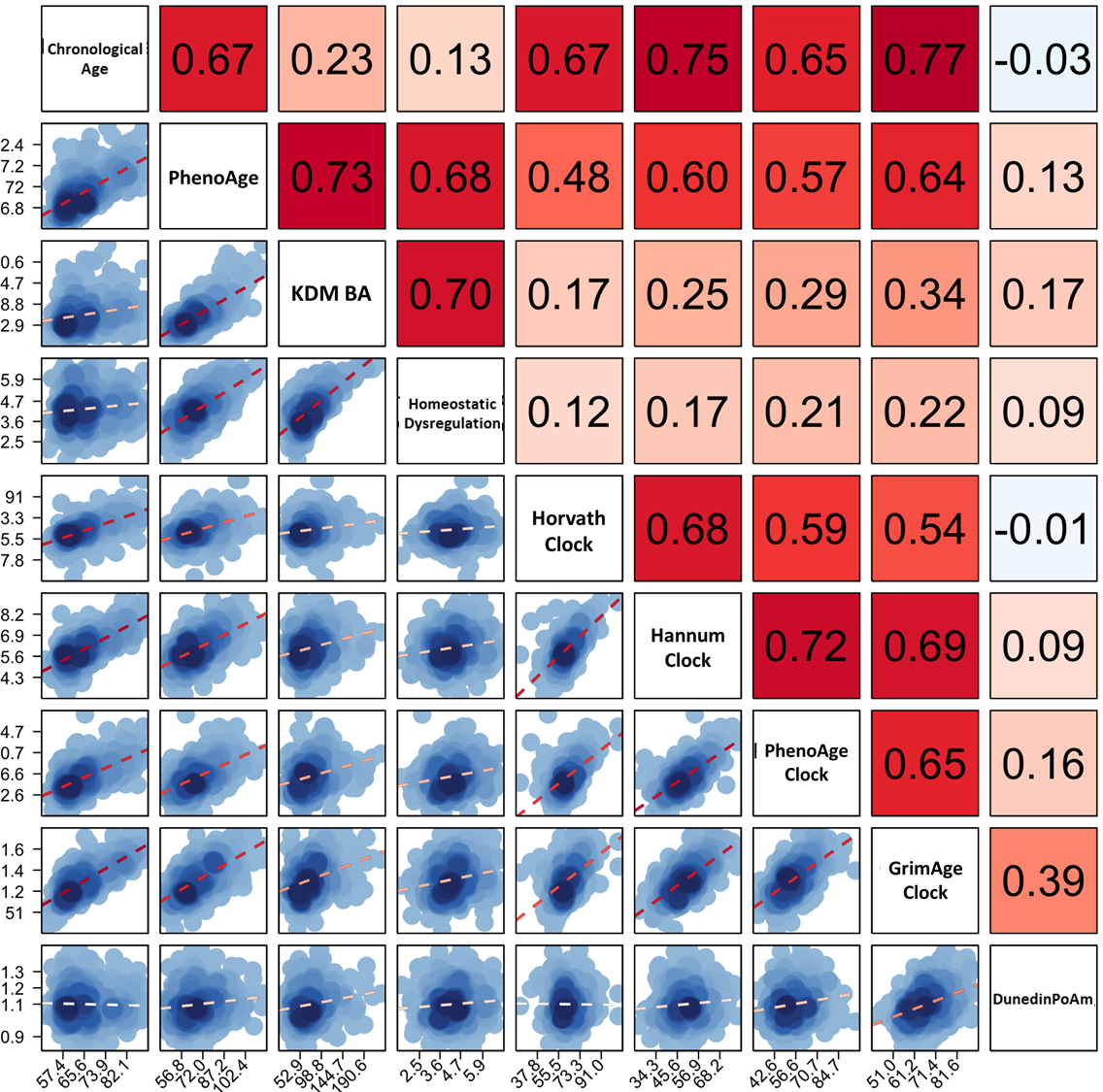

**Supplemental Figure S2. Associations of biological aging measures with healthspan characteristics in Black and White US Health and Retirement Study participants.** The figure graphs associations of biological aging measures with healthspan characteristics estimated from stratified samples of Black and White participants. Sample sizes are reported in the Y-axis label. The Y-axis shows the predicted count of functional impairments, ADLs, and chronic conditions. The X-axis shows biological aging, denominated in SD units.

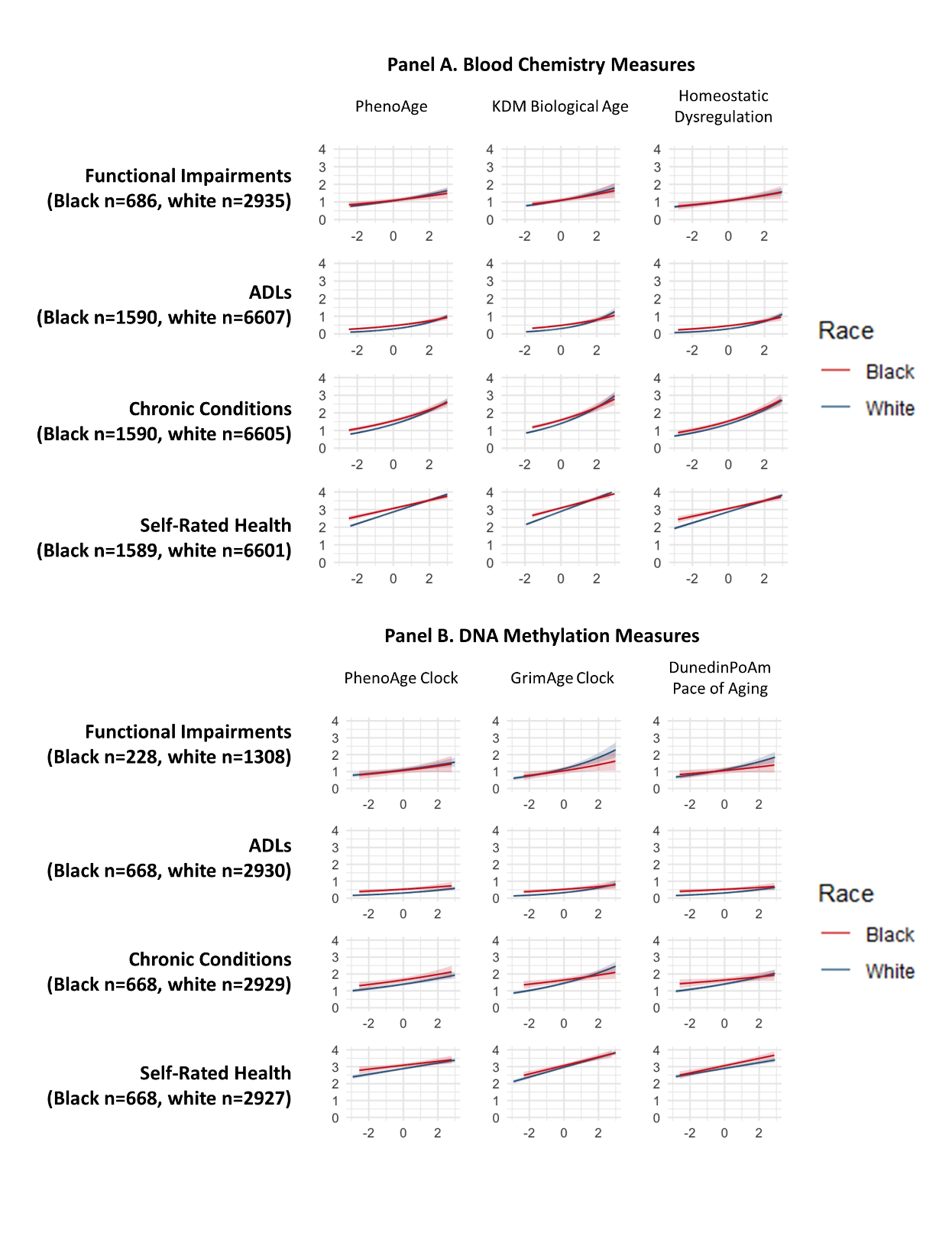

**Supplemental Figure S4. Distribution of prevalent (2016) healthspan-related characteristics in the Venous Blood Study and DNA-methylation HRS subsamples.** This figure graphs the distribution of functional impairments, activities of daily living limitations, physician-diagnosed chronic conditions, and self-rated health exhibited/reported by participants in the sample. Over a two-year follow-up period, 10% of the white VBS sample (n=608) and 12% of white DNA-methylation sample (n=297) developed a new limitation in performing activities of daily living, compared to 13% of the Black VBS sample (n=184) and 11% of the Black DNA-methylation sample (n=66). Over the same period, 13% of the white VBS sample (n=766) and 14% of white DNA-methylation sample (n=357) developed a new chronic condition, compared to 11% of the Black VBS sample (n=157) and 10% of the Black DNA-methylation sample (n=60). 23% of the white VBS sample (n=1325) and 23% of white DNA-methylation sample (n=583) developed a new chronic condition, compared to 24% of the Black VBS sample (n=335) and 24% of the Black DNA-methylation sample (n=142). 2.9% of the white VBS sample (n=194) and 3.5% of white DNA-methylation sample (n=102) developed a new chronic condition, compared to 2.7% of the Black VBS sample (n=43) and 3.1% of the Black DNA-methylation sample (n=21).

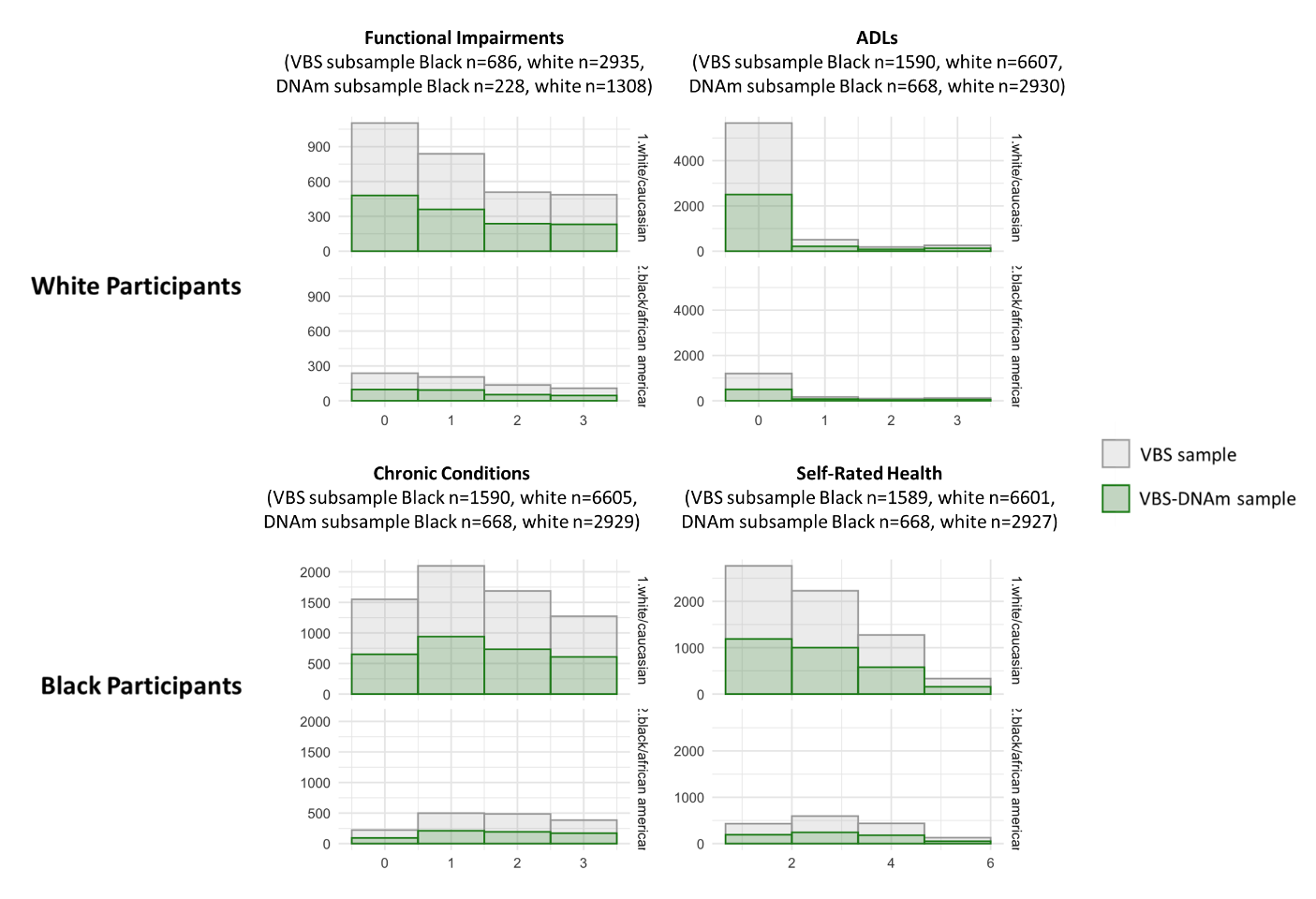

### Supplemental Tables

**Supplemental Table S1. Demographic characteristics of HRS sample, VBS subsample, and VBS-DNAm subsample.** Full HRS sample consists of all White and Black participants in the 2016 Health and Retirement study (n=29192). Full VBS sample consists of all White and Black participants in the 2016 Health and Retirement study who also provided biomarker data via participation in the Venous Blood Study (n=8198). VBS-DNA Methylation Sample consists of all White and Black participants in the 2016 Health and Retirement study who provided DNA-methylation data via participation in the Venous Blood Study (n=3598). Biological-age residuals were calculated by fitting a regression of biological age on chronological age to the full VBS-DNA Methylation Sample, then subtracting the fitted value from that estimated using blood-chemistry algorithms or DNA-methylation clock calculations.

*See Supplemental Excel Table File.*

**Supplemental Table S2a. Effect-sizes for cross-sectional associations of biological-aging measures with healthspan-related characteristics.** The table shows effect-sizes for associations of biological-aging measures derived from blood chemistries (Panel A), DNA-methylation (Panel B), and chronological age (Panel C) with healthspan-related characteristics. For functional impairments, ADLs, and chronic conditions, effect-sizes are incidence rate ratios (IRRs) estimated from Poisson regression for 1-SD increments of the biological aging measures. For self-rated health, effect-sizes are Pearson's r correlations estimated from linear regression for 1-SD increments of the biological aging measures. SDs were computed from the full samples for which each measure of biological aging was available (see Supplemental Table S1). Full sample estimates are weighted using Venous Blood Study and DNA-methylation sample weights provided by HRS and adjusted for age, and sex. Race-stratified estimates are unweighted and adjusted for age, sex, and census region of residence. Sample sizes for VBS/VBS-DNAm samples are n=3721/1676 for functional impairment; n=8484/3785 for ADLs; n=8482/3784 for chronic conditions; n=8476/3782 for self-rated health. The sample size for analysis of the functional impairment index is smaller because only ~50% of 2016 Venous Blood Study participants were also included in the in-person assessments of physical function conducted in that wave of data collection. The table illustrates three findings. First, faster/more-advanced biological aging predicts decline in healthspan. Second, effect-sizes tended to be smaller for analysis of Black as compared to White HRS participants. Third, Black-White differences in effect-sizes were parallel for analysis of biological aging measures and of chronological age.

*See Supplemental Excel Table File.*

**Supplemental Table S2b. Effect-sizes for longitudinal associations of biological-aging measures with healthspan-related characteristics.** The table shows effect-sizes for biological-aging measures derived from blood chemistries (Panel A) and DNA-methylation (Panel B), and chronological age (Panel C). For mortality, effect-sizes are hazard ratios estimated from Cox regression. For incident ADLs and incident chronic conditions, effect-sizes are incidence rate ratios (IRRs) estimated from Poisson regression. For changes in self-rated health, effect-sizes are standardized regression coefficients (interpretable as Pearson r) estimated from linear regression. All effect-sizes are reported for 1-SD increments of the biological aging measures. Full sample estimates are weighted using Venous Blood Study and DNA-methylation sample weights provided by HRS and adjusted for age, and sex. Race-stratified estimates are unweighted and adjusted for age, sex, and census region of residence. Sample sizes for VBS/VBS-DNAm samples are n=8484/3785 for mortality; n=7491/3327 for ADLs; n=7497/3329 for chronic conditions; n=7488/3326 for self-rated health. The sample size for analysis of the functional impairment index is smaller because only ~50% of 2016 Venous Blood Study participants were also included in the in-person assessments of physical function conducted in that wave of data collection. The table illustrates three findings. First, more-advanced/faster biological aging at baseline is associated with mortality, incident disability and disease, and declines in self-rated health over 2-3 years of follow-up. Second, effect-sizes tended to be smaller for analysis of Black- as compared to White-identifying HRS participants. Third, Black-White differences in effect-sizes for analysis of biological aging were paralleled in analysis of chronological age.

*See Supplemental Excel Table File.*

**Supplemental Table S3. Effect-sizes for Black-White disparities in healthspan-related characteristics before and after covariate adjustment for measures of biological aging.** The table shows regression estimates of Black-White disparities in longitudinal change in healthspan-related characteristics, before and after statistical adjustment for biological aging measures. Estimates are interpretable as incident rate ratios (IRRs) for functional impairments, ADLs, and chronic conditions, as Cohen's D for self-rated health, and as hazard ratios (HRs) for mortality. Unadjusted estimates of disparities are calculated by regressing healthspan characteristics on Black vs. White race with covariate adjustment for age, sex, and census region of residence. Adjusted estimates are calculated by adding a measure of biological aging to the base model. The table shows that adjustment for biological aging measures attenuates effect-sizes by 20-30% across outcomes.

*See Supplemental Excel Table File.*

**Supplemental Table S4a. Tests of biological aging as a mediator of Black-White differences in prevalent healthspan characteristics in models with and without exposure-mediator interactions.** Table shows results of mediational analysis using the approach of Vanderweele (2014). For each healthspan characteristic, there are two columns of estimates and 95% Confidence Intervals. The first columns show results from mediational models without exposure-mediator interactions. The second columns show results with exposure-mediator interactions. The first table row shows the controlled direct effect (CDE), second row shows the pure natural direct effect (PNDE), third row shows the total natural direct effect (TNDE), fourth row shows the pure natural indirect effect (PNDE), fifth row shows the total natural indirect effect (TNIE), sixth row shows the total effect (TE), last row shows the proportion mediated (PM). For estimates of the controlled direct effect, the value of the mediator (biological-age) is set to zero. Because our biological aging measures are transformed to a Z scaling (M=0, SD=1), a zero value of the mediator corresponds to the extent or pace of biological aging that is at the sample norm. Note that the total effect is equal to the sum of the pure natural direct effect and the total natural indirect effect (PNDE + TNIE), and to the sum of the total natural direct effect and the pure natural indirect effect (TNDE + PNIE).

*See Supplemental Excel Table File.*

**Supplemental Table S4b. Tests of biological aging as a mediator of Black-White differences in incident healthspan characteristics in models with and without exposure-mediator interactions.** Table shows results of mediational analysis using the approach of Vanderweele (2014). For each healthspan characteristic, there are two columns of estimates and 95% Confidence Intervals. The first columns show results from mediational models without exposure-mediator interactions. The second columns show results with exposure-mediator interactions. The first table row shows the controlled direct effect (CDE), second row shows the pure natural direct effect (PNDE), third row shows the total natural direct effect (TNDE), fourth row shows the pure natural indirect effect (PNDE), fifth row shows the total natural indirect effect (TNIE), sixth row shows the total effect (TE), last row shows the proportion mediated (PM). For estimates of the controlled direct effect, the value of the mediator (biological-age) is set to zero. Because our biological aging measures are transformed to a Z scaling (M=0, SD=1), a zero value of the mediator corresponds to the extent or pace of biological aging that is at the sample norm. Note that the total effect is equal to the sum of the pure natural direct effect and the total natural indirect effect (PNDE + TNIE), and to the sum of the total natural direct effect and the pure natural indirect effect (TNDE + PNIE).

*See Supplemental Excel Table File.*

**Supplemental Table S5. Tests of Black-White differences in biological aging associations with healthspan characteristics.** The table shows results from tests of super-multiplicative and super-additive interaction. For the functional impairment, ADL, chronic conditions, and mortality outcomes, super-multiplicative interaction is tested by the product term for racial identity*biological aging measure in the regression of the healthspan outcome on biological aging, racial identity, their product term, age, sex, and census region. The product-term coefficient tests the hypothesis that biological aging-measure associations with the healthspan outcome differ between Black and White participants on a multiplicative scale. Super-additive interaction is tested in Relative Excess Risk due to Interaction (RERI) analysis of the same model. The RERI tests differences in association on an additive scale. Confidence intervals for RERIs were estimated using the bootstrap method. For self-rated health, the product term from linear regression tests the hypothesis that biological age associations differ between Black and White participants on an additive scale. For comparison, results from models that replace biological aging measures with chronological age are reported at the bottom of the table.

*See Supplemental Excel Table File.*
